# Computational modeling enables individual assessment of postprandial glucose and insulin responses after bariatric surgery

**DOI:** 10.1101/2024.11.25.24317927

**Authors:** Onur Poyraz, Sini Heinonen, S. T. John, Tuure Saarinen, Anne Juuti, Pekka Marttinen, Kirsi H. Pietiläinen

## Abstract

Bariatric surgery enhances glucose metabolism, yet the detailed postprandial joint glucose and insulin responses, variability in individual outcomes, and differences in surgical approaches remain poorly understood. To address this, we used hierarchical multi-output Gaussian process (HMOGP) regression to reveal clinically relevant patterns between persons undergoing two types of bariatric surgery by modeling the individual postprandial glucose and insulin responses and estimating the average response curves from individual data. 44 participants with obesity underwent either Roux-en-Y gastric bypass (RYGB; n=24) or One-Anastomosis gastric bypass (OAGB; n=20) surgery. The participants were followed up at the 6th and 12th months after the operation, during which they underwent an oral glucose tolerance test (OGTT) and a mixed meal test (MMT). A marked reduction in glycemia, an earlier glucose peak, and an increase and sharpening in the postprandial glucose and insulin responses were evident in both metabolic tests post-operation. MMT resulted in higher postprandial glucose and insulin peaks compared with OGTT. Higher glucose and insulin responses were observed after RYGB compared with OAGB, suggesting differences between the procedures that may influence the clinical practice. Computational modeling with HMOGP regression can thus be used to, in detail, predict the combined responses of patient cohorts to ingested glucose or a mixed meal and help in assessing individual metabolic improvement after weight loss. This can lead to new knowledge in personalized metabolic interventions.

## 1 Introduction

The prevalence of obesity has increased enormously over the last decades. Over 1.8 billion people lived with overweight or obesity in 2017 (WHO factsheet). Obesity is associated with various diseases and metabolic problems such as type 2 diabetes (T2DM), dyslipidemia, cardiovascular diseases, and cancer (Zimmet et al., 2005; Guh et al., 2009).

Treating obesity is extremely difficult. While diet-induced weight loss strategies lead to modest results, bariatric surgery is the most effective treatment of obesity, leading to improved glucose tolerance already within days (Adams et al., 2012; Sjöström, 2013). Roux-en-Y gastric bypass (RYGB) has been the gold standard in bariatric surgery. One-Anastomosis gastric bypass (OAGB) emerged as a potentially more beneficial procedure for the resolution of T2DM (Almalki et al., 2018; Magouliotis et al., 2018), but recent randomized controlled trials have challenged this (Disse et al., 2014; Robert et al., 2019; Heinonen et al., 2023). Additionally, individual metabolic phenotypes of participants undergoing bariatric surgery, such as differential glucose response between sexes (Mauvais-Jarvis, 2018), have resulted in the need for understanding and modeling in detail the glycemic responses in different patient cohorts before and after surgery.

Conventional methods for assessing glucose responses after the oral glucose tolerance test (OGTT) and the mixed meal test (MMT) have relied on averaging the means of patient responses for each individual separately. Previously used methods in assessing the results of these tests therefore yield a robust estimation of blood glucose regulation but lack detailed characteristics of the overall blood glucose/insulin response at the population level and neglect individual differences. Additionally, OGTT is sparsely sampled and individual estimates may be missing from the available data. Artificial intelligence techniques to better understand blood glucose regulation have emerged in recent years (Li et al., 2020). For example, the Eindhoven Diabetes Education Simulator (E-DES) model has been successfully used for OGTT to develop personalized models of glucose and insulin sensitivity and to explore the heterogeneity in the responses (Erdős et al., 2021). In another study, methods quantifying glucose fluxes between tissues using tracers have been applied to create predictive models of glucose-insulin interactions in MMT analyses (Dalla Man et al., 2007). Zeevi et al. (2015) developed a machine-learning algorithm integrating blood parameters, dietary habits, anthropometrics, physical activity, and gut microbiota to estimate the glucose and insulin responses accurately. A Gaussian process has been previously used in modeling OGTT responses in participants with and without cystic fibrosis to provide measures of beta-cell function with quantified uncertainty (Garrish et al., 2023).

In this study, we developed hierarchical multi-output Gaussian process (HMOGP) regression, which is the combination of hierarchical Gaussian process (HGP) and multi-output Gaussian process (MOGP). The hierarchical Gaussian process (HGP) is beneficial when part of the data reveals different characteristics, such as the considerable inter-individual variability usually observed in postprandial glucose and insulin responses (Park and Choi, 2010). On the other hand, the multi-output Gaussian process (MOGP) is able to deal with multiple correlated outputs (such as glucose and insulin dynamics) and to provide more accurate predictions than simply modeling these outputs separately (Álvarez et al., 2010; Liu et al., 2018; Moreno-Muñoz et al., 2018). Similar techniques to HMOGP have been successfully applied before in different domains such as anomaly detection (Cho et al., 2019), multi-task learning (Li and Chen, 2018), and mobile cellular network analysis (Cuesta Ramírez, 2015), performing better than MOGP and GP. We expanded these techniques to the medical domain in OGTT and MMT responses with necessary adaptations. HMOGP allows us (1) to model the insulin and glucose response together via multi-output (2) while considering the individual level differences through a hierarchical structure. It also (3) quantifies uncertainty in predictions, making it suitable for datasets with noise or missing values, and (4) can model non-linearities and complex temporal dynamics in postprandial responses, providing more accurate predictions than simpler models (Williams and Rasmussen, 2006). Modeling both OGTT and MMT data in detail with HMOGP regression allows for a more detailed assessment of postprandial glucose and insulin responses. We evaluated model outcomes to better understand the postprandial glucose and insulin levels after an OGTT and an MMT, considering before and after different types of bariatric surgery and between sexes or participants with or without T2DM, providing a thorough analysis of postprandial response with an advanced technique.

## 2 Methods

### 2.1 Subjects

We included participants with obesity (n=44, aged 46.6 years with 14 men and 30 women, 26 participants with obesity, 18 participants with obesity and T2DM, matched for age and diabetes status between the operation types). Participants were examined before and at 6th and 12th months after bariatric surgery with either Roux-en-Y gastric bypass (RYGB; n=24, 8 men, 16 women, n=9 with T2DM) or One-Anastomosis gastric bypass (OAGB; n=20, 6 men, 14 women, n=9 with T2DM). 33% of men vs. 43% of women had T2DM. The subjects were recruited from Helsinki University Hospital through the Department of Gastrointestinal Surgery. The full randomized clinical trial is registered at clinicaltrials.gov with no. NCT02882685 and the randomization and selection process of the participants and study dropouts are described in detail in Saarinen et al. (2019) and Heinonen et al. (2023). All participants from Helsinki University Hospital with OGTT data available were included in the current study. No data or participants were removed from the analysis. Written informed consent was obtained from all participants. The study protocol was designed and performed according to the principles of the Helsinki Declaration and approved by the Ethical Committee of the Helsinki University Central Hospital.

### 2.2 Bariatric surgery

In RYGB, the gastric pouch was created with one horizontal 45 mm and two vertical 60 mm staplers. The length of the biliary limb was 80 cm, and that of the alimentary limb was 130 cm. In OAGB, a tubular gastric pouch was created using 60 mm staplers along a 38Fr bougie starting at the crow’s foot with a horizontal 45 mm stapler and the omega loop being 210 cm long. The length of the bypasses was standardized between the procedures to allow for equal comparison. A 210 cm biliopancreatic limb in OAGB and 80 cm biliopancreatic and 130 cm alimentary limbs in RYGB were chosen to obtain equally long bypassed intestines in both groups (Saarinen et al., 2019).

### 2.3 Clinical examinations and body composition

Weight and height were measured after an overnight fast in light clothing. Whole body composition was measured by Dual-energy X-ray absorptiometry (DEXA) using a Lunar Prodigy whole-body scanner (GE Medical Systems, Madison, WI).

### 2.4 Analytical blood samples

Fasting laboratory tests, including Hba1c and plasma lipids, were performed as described by Heinonen et al. (2015).

### 2.5 Mixed meal test (MMT) and oral glucose tolerance test (OGTT)

Glucose metabolism was measured after an overnight fast with a 3-hour oral glucose tolerance test (OGTT, 75 g of glucose), with time points 0 (before the ingestion of the glucose drink) and post-glucose samples at 30, 60, 120, and 180 min. We additionally performed a 6-hour Mixed Meal Test (MMT) on a separate day, where a fasting blood sample was collected before ingesting the liquid meal of 2620 kJ (627 kcal) with a balanced distribution of fat (24 g), carbohydrates (76 g) and protein (24 g) (Resource® 2.5 Compact, Nestle Health Science), with post-meal samples at 15, 30, 60, 120, 180, 240 and 360 min for measuring glucose and insulin.

### 2.6 Data features

Each patient underwent both OGTT and MMT tests across three different visits: once preoperatively and then at the 6th and 12th months postoperatively. Six participants missed the baseline OGTT visit, while 10 and 6 missed the 6th and 12th month visits, respectively. In MMT, 4 and 5 participants missed the 6th and 12th month visits, respectively. Additionally, 3% of the overall data was missing due to vomiting or nausea, which interrupted the test.

### 2.7 Statistical method

Blood glucose and insulin evolve as a function of time. Here, we model them using a Gaussian process, which is a continuous stochastic process and can be considered a probability distribution over functions (MacKay et al., 1998; Williams and Rasmussen, 2006). The standard Gaussian process regression is defined as:

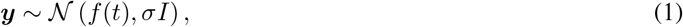

where ***y*** *∈*ℝ ^*T*^ are the observation points, *t ∈{*1, …, *T}* the input locations, and *σ* the measurement error. The function *f* (*t*) is defined as the Gaussian process *f* (*t*) *~ 𝒢 𝒫* (0, **K**) with kernel matrix **K**. However, applying this approach directly to our problem is not straightforward. First, we know that glucose and insulin responses are strongly correlated, and applying Gaussian process regression on them separately would miss this correlation. Therefore, we propose to use Multi-Output Gaussian Process regression (Álvarez et al., 2010; Liu et al., 2018; Moreno-Muñoz et al., 2018), which is able to calculate the cross-covariance between functions and is the proper choice for modeling insulin and glucose together. If we define ***f*** = [*f*_glucose_, *f*_insulin_]:

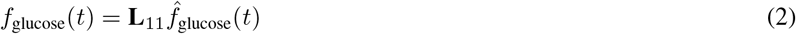

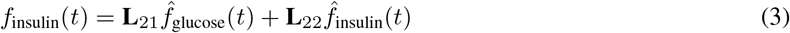

where 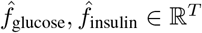 are independent GPs and **L** is the Cholesky factor matrix of the corresponding correlation matrix **C**, then ***f*** *∈* ℝ ^*T×D*^, where D is the output dimension (glucose, insulin), has the multi-output GP distribution. However, the challenge of the data is not limited to the output correlation. The second issue is that individuals have slightly different blood glucose and insulin responses. Hierarchical Gaussian process regression modeling can solve this issue by learning the population average response curve and individual deviations from the average response separately (Park and Choi, 2010). Let ***f*** _*n*_(*t*) denote the response curve for a single patient, ***f*** ^population^(*t*) the average response curve across all participants, and 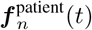 the deviation of the patient from the average response. Then, hierarchical GP can be represented as:

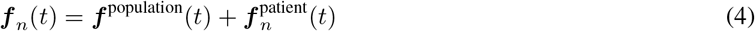

where 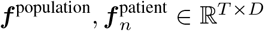 have multi-output Gaussian process distributions. We apply the same hierarchical approach to model the constant terms for fasting glucose and insulin, as follows:

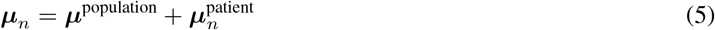

where ***µ***_*n*_ is fasting glucose and insulin for a single patient, ***µ***^population^ corresponds to the normally distributed population average of fasting glucose and insulin, and 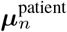denotes the normally distributed individual deviation from the population average.

Overall, we modeled the data using the Hierarchical Multi-Output Gaussian process (HMOGP), where hierarchy allows us to consider the deviations of individual responses, and multi-output models the correlation of insulin and glucose response. Let the whole data be denoted by ***y*** *∈* ℝ ^*N×T×D*^, where *N* is the total number of participants, *T* is the number of time points and *D* = 2 refers to the two possible outputs (glucose, insulin). If we index patient by *n*, time by *t*, and output by *d*, the blood glucose and insulin level ***y***_*n,t*_ of patient *n* at time *t* can be represented as

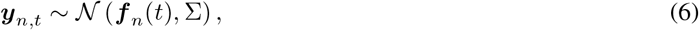

where Σ is the diagonal measurement error matrix, and the latent function ***f*** _*n*_ is formulated as

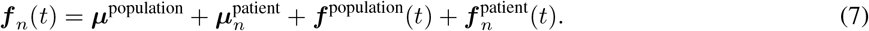

We applied this model separately to the different visits (Baseline, 6th month, 12th month) and tests (OGTT, MMT). The only shared parameter across these models was the measurement error. For comparisons by operation, sex, and T2DM, we divided the dataset accordingly and modeled them separately. Further details of the model parameters and priors, along with a link to the Stan code of the model, can be found in Appendix A.

Stan software (Carpenter et al., 2017) was used to draw samples from the posterior distributions using the Markov Chain Monte Carlo inference. We used posterior samples to calculate Bayesian credible intervals (CI) and *p*-values as described in Gelman et al. (2013). Furthermore, we evaluated four metrics to investigate the characteristics of the postprandial glucose and insulin responses: peak value (PV), time-normalized area under the curve (AUC), peaking time (PT), and time in the risk zone (TiH; Hyperglycemia for glucose *>* 7.0mmol/l and Hyperinsulinemia for insulin *>* 50mU/l). These metrics are illustrated in Figure 1. In the clinical analyses, non-normally distributed variables were log10 transformed before the parametric analyses. Differences at baseline between the groups were analyzed by Student’s T-tests and between visits with generalized linear mixed modeling adjusting for sex, T2DM, and operation type (Stata Statistical software ver. 7.0). These results are presented as means and 95% confidence intervals (CI).

**Figure 1:**
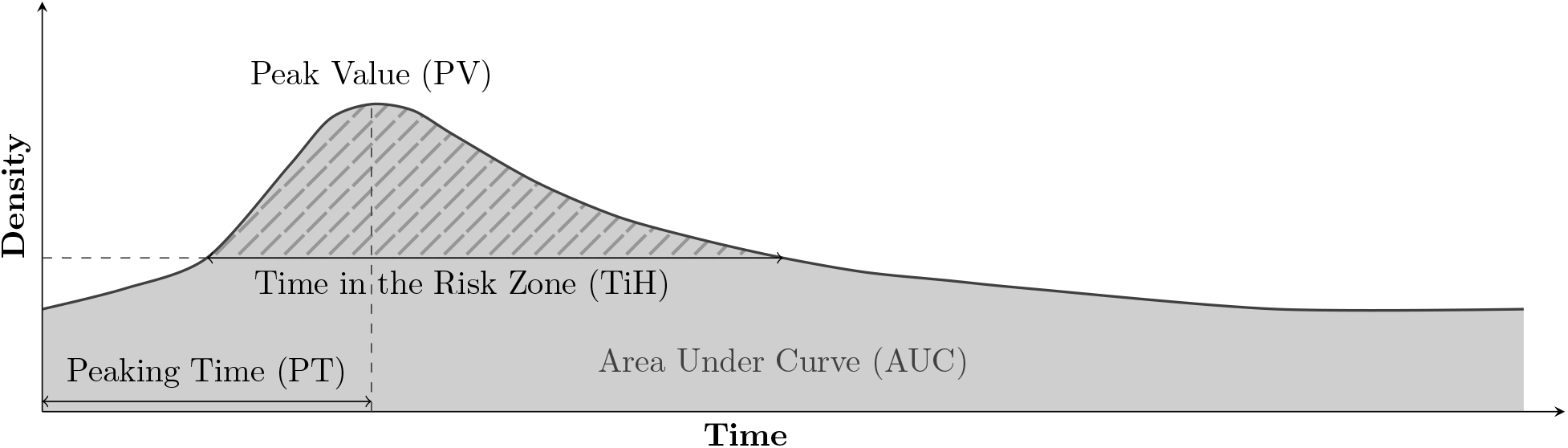
Definition of the metrics evaluated in the experiments. Peaking Time (PT) and Time in the Risk Zone (TiH; Hyperglycemia for glucose > 7.0mmol/l and Hyperinsulinemia for insulin > 50mU/l) represented in minutes, and Peak Value (PV) and time-normalized Area Under Curve (AUC) represented in mmol/l for glucose and mU/l for insulin.

## 3 Results

Throughout the manuscript, we investigated the glucose and insulin responses of all participants following an oral glucose tolerance test (OGTT) and a mixed meal test (MMT) over the entire dataset and in different subgroups divided by operation, sex, and T2DM using hierarchical multi-output Gaussian process (HMOGP) regression, which is suitable for longitudinal data. In MMT, 76g of glucose was combined with protein and fats, while OGTT had only 75g of glucose. Figure B.1 and Figure B.2 show HMOGP predictions by individuals, which validates the model fit for different persons. Note that Figure B.1 and Figure B.2 show only the participants with the complete set of observations while the model was fitted using all the participants.

### 3.1 Glucose and insulin responses sharpen post-surgery in OGTT and MMT

From baseline to the 12th month of post-surgery, the glucose-insulin response improved in both tests (Figure 2). During OGTT, AUCs of glucose and insulin concentrations significantly decreased from baseline to 6th and 12th months (Table F.3). Similarly, PVs of both glucose and insulin were reduced significantly from baseline to the 12th month (Table F.2), and PT at the 12th month occurred 12 minutes earlier for glucose and 11 minutes earlier for insulin compared to baseline (Table F.1). TiH for glucose was reduced by 53 min during weight loss, already bringing about large metabolic benefits (Table F.4). Overall, OGTT demonstrated a sharpened and significantly earlier glucose and insulin response with lower peak values following bariatric surgery (Figure 2).

**Figure 2:**
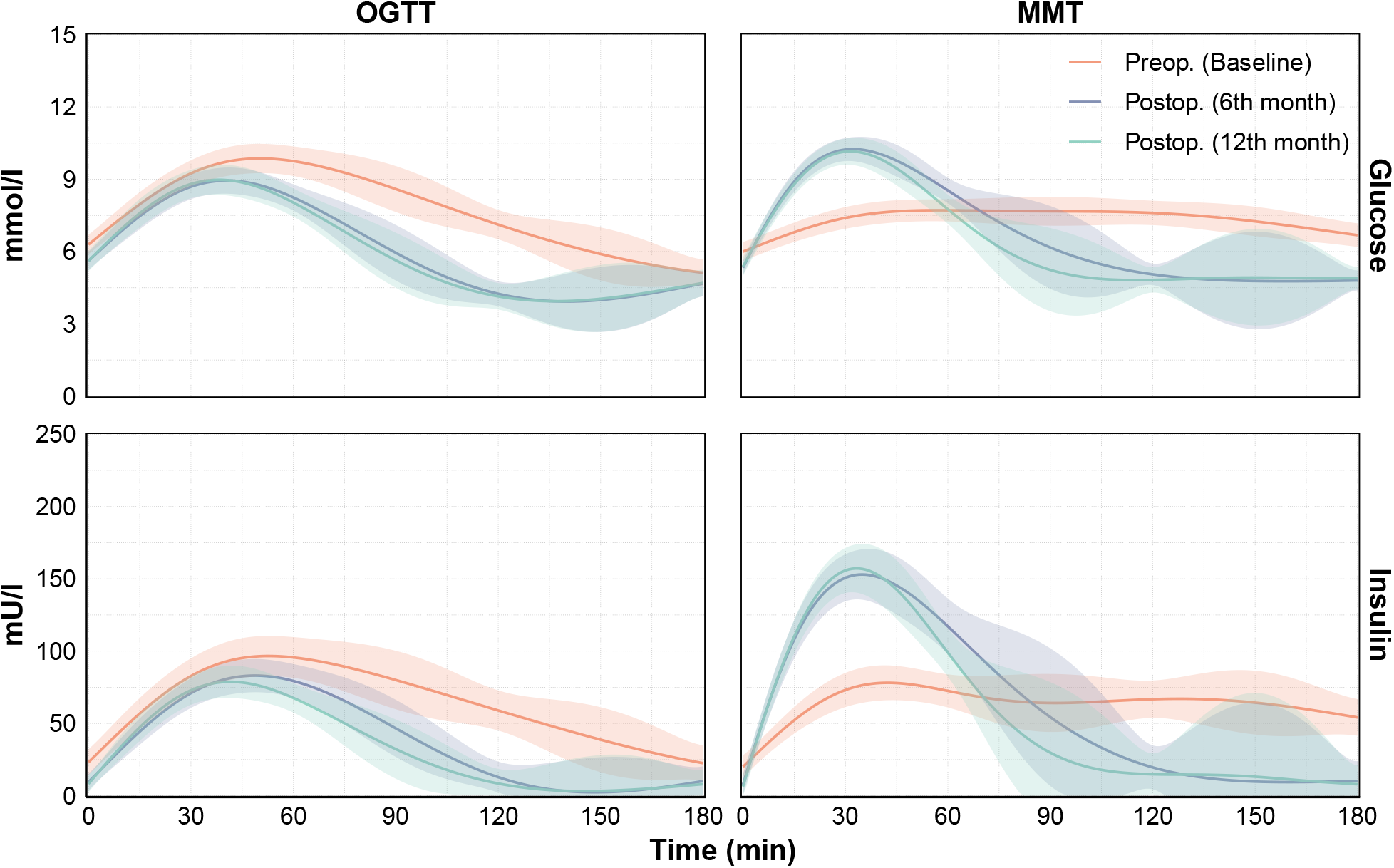
The comparison of model predicted average glucose and insulin responses in different visits (Baseline, 6th month, and 12th month) within OGTT and MMT tests. Shaded areas represent 95% confidence intervals (CI).

In MMT, AUCs of both glucose and insulin showed a significant reduction from baseline to the 12th month (Table F.3). Unlike in OGTT, PVs of glucose and insulin significantly increased over 12 months (Table F.2). PT occurred 48 minutes and 11 minutes earlier for glucose and insulin, respectively, from baseline to the 12th month (Table F.1). Additionally, there was a total reduction of 83 minutes in TiH for glucose from baseline to the 12th month (Table F.4). These findings indicate a sharper and more pronounced glucose-insulin response, with significantly higher peak values and earlier peak times after bariatric surgery in MMT (Figure 2).

### 3.2 Higher postprandial glucose-insulin response in MMT vs. OGTT after bariatric surgery

OGTT and MMT evaluate participants’ metabolic status through differential ingestion of macro-nutrients between the tests. To elucidate the differences between the tests before and after bariatric surgery, we next analyzed the average postprandial response in glucose and insulin, comparing MMT and OGTT by including similar follow-up times of 180 minutes in both tests (Figure 3). PVs of glucose and insulin were higher after OGTT than MMT at baseline, but other metrics did not differ significantly at baseline (Appendix F). Post-operatively at the 6th and 12th months, glucose and insulin responses (AUC, PV, and PT) were all significantly enhanced in MMT compared with OGTT (Appendix F). TiH for glucose did not differ between the tests during weight loss. While both tests are performed in the same individuals and on following days, these differences could be attributed to differential absorption of glucose in combination with other macro-nutrients in MMT vs. OGTT post-operatively and to postprandially enhanced insulin stimulus originating from proteins and fats included in MMT.

**Figure 3:**
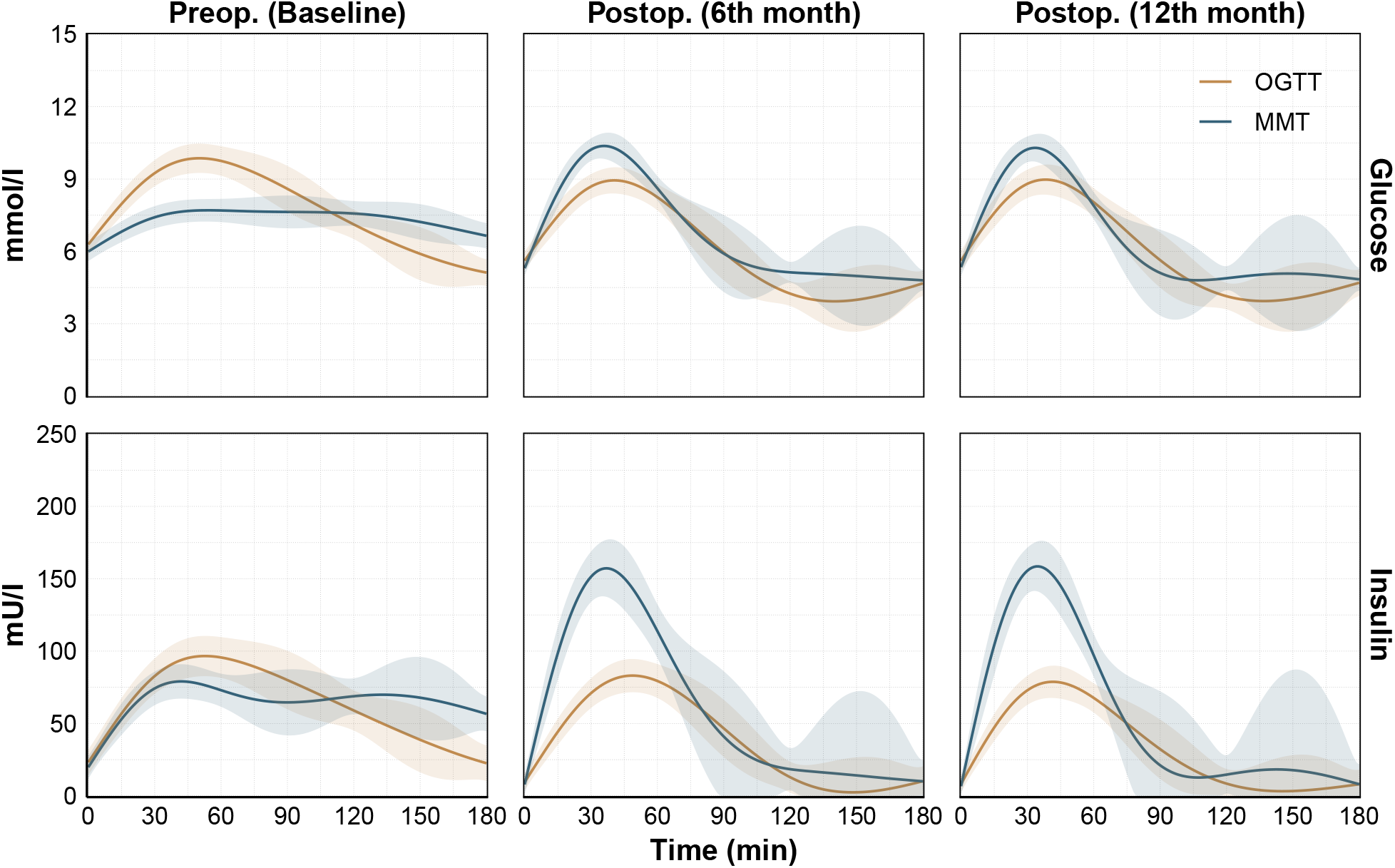
The comparison of model predicted average glucose and insulin responses by OGTT and MMT test in different visits. Shaded areas represent 95% confidence intervals (CI).

### 3.3 Glucose-insulin response is higher and longer after RYGB than OAGB

To shed light on the potential differential responses between two types of bariatric surgery, we modeled RYGB (n=24) and OAGB (n=20) groups separately. Figure 4 and Figure 5 show population comparison for OGTT and MMT tests, respectively. In OGTT, the RYGB and OAGB groups had comparable glucose and insulin responses at baseline. The RYGB group showed a trend towards a larger AUC of glucose compared to the OAGB group (p=0.09) at the 12th month and had a significantly larger AUC of insulin at the 6th month (Table F.3). PV of glucose for the RYGB group was higher than the OAGB group at the 12th month (10.0 mmol/l vs. 7.8 mmol/l). For insulin, the RYGB group had a higher PV at the 6th month and a trend towards it at the 12th month (p=0.062, Table F.2). PTs showed no difference between operations (Table F.1)). TiH for glucose was longer in RYGB vs. OAGB group at the 12th month (70 min vs. 47 min), but this difference was nearly significant already at baseline (125 min vs. 105 min, p=0.088, Table F.4). During the entire weight-loss period, the mean TiH reduction of glucose was similar between RYGB and OAGB (55 min vs. 58 min).

**Figure 4:**
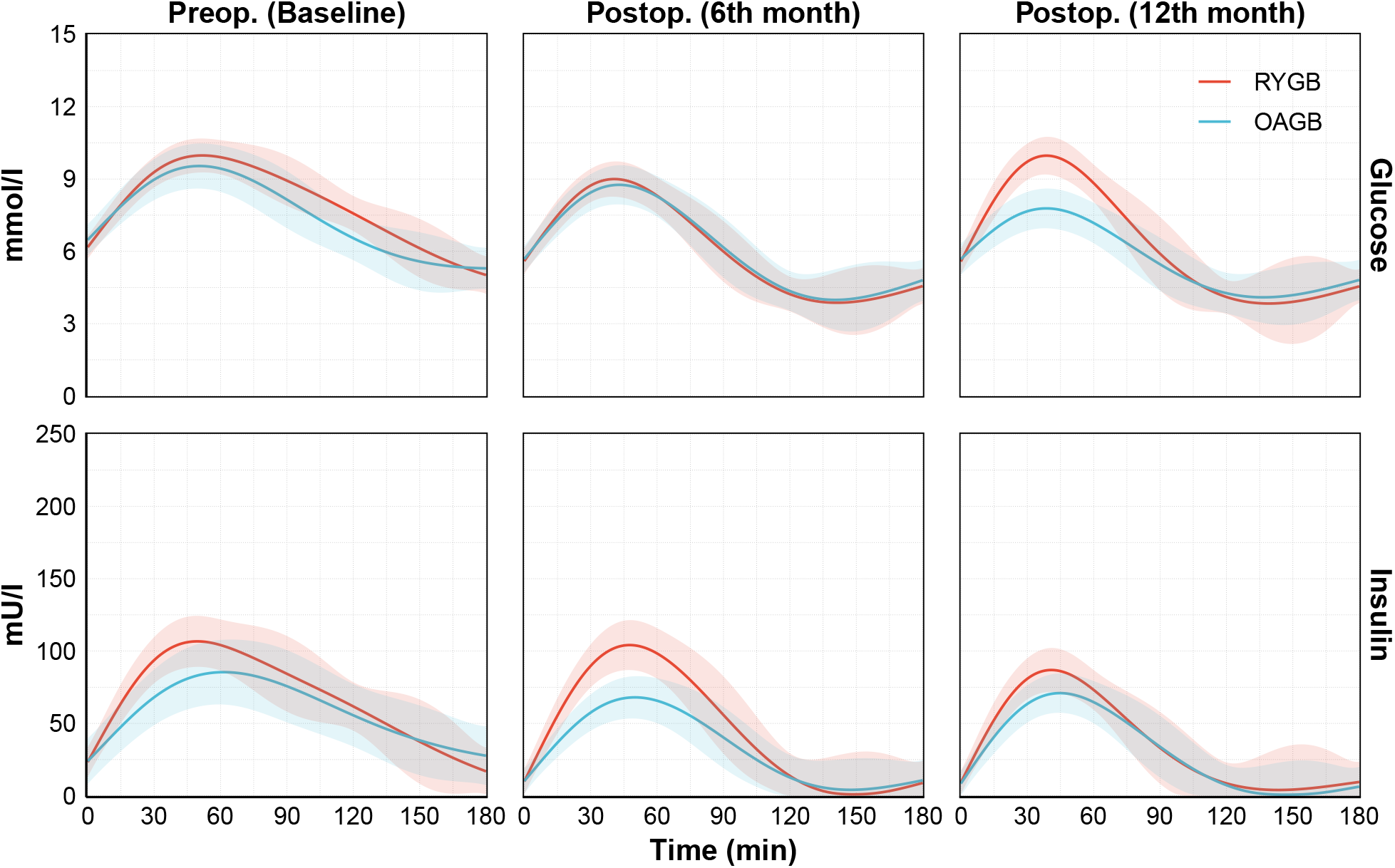
The comparison of model predicted average glucose and insulin response of the participants with Roux-en-Y gastric bypass (RYGB) and One-Anastomosis gastric bypass (OAGB) operation in OGTT test. Shaded areas represent 95% confidence intervals (CI).

**Figure 5:**
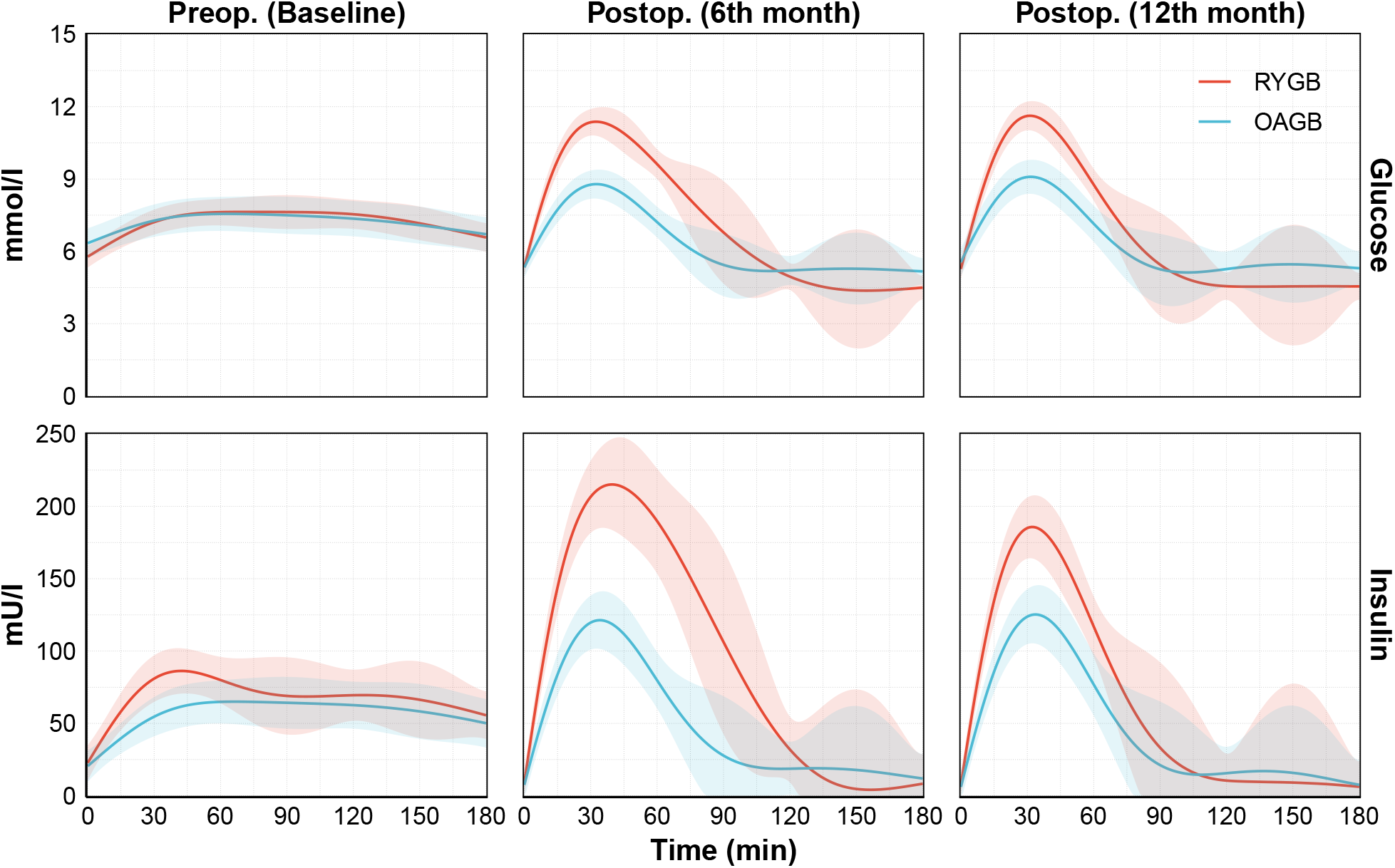
The comparison of model predicted average glucose and insulin response of the participants with Roux-en-Y gastric bypass (RYGB) and One-Anastomosis gastric bypass (OAGB) operation in MMT test. Shaded areas represent 95% confidence intervals (CI).

**Figure 6:**
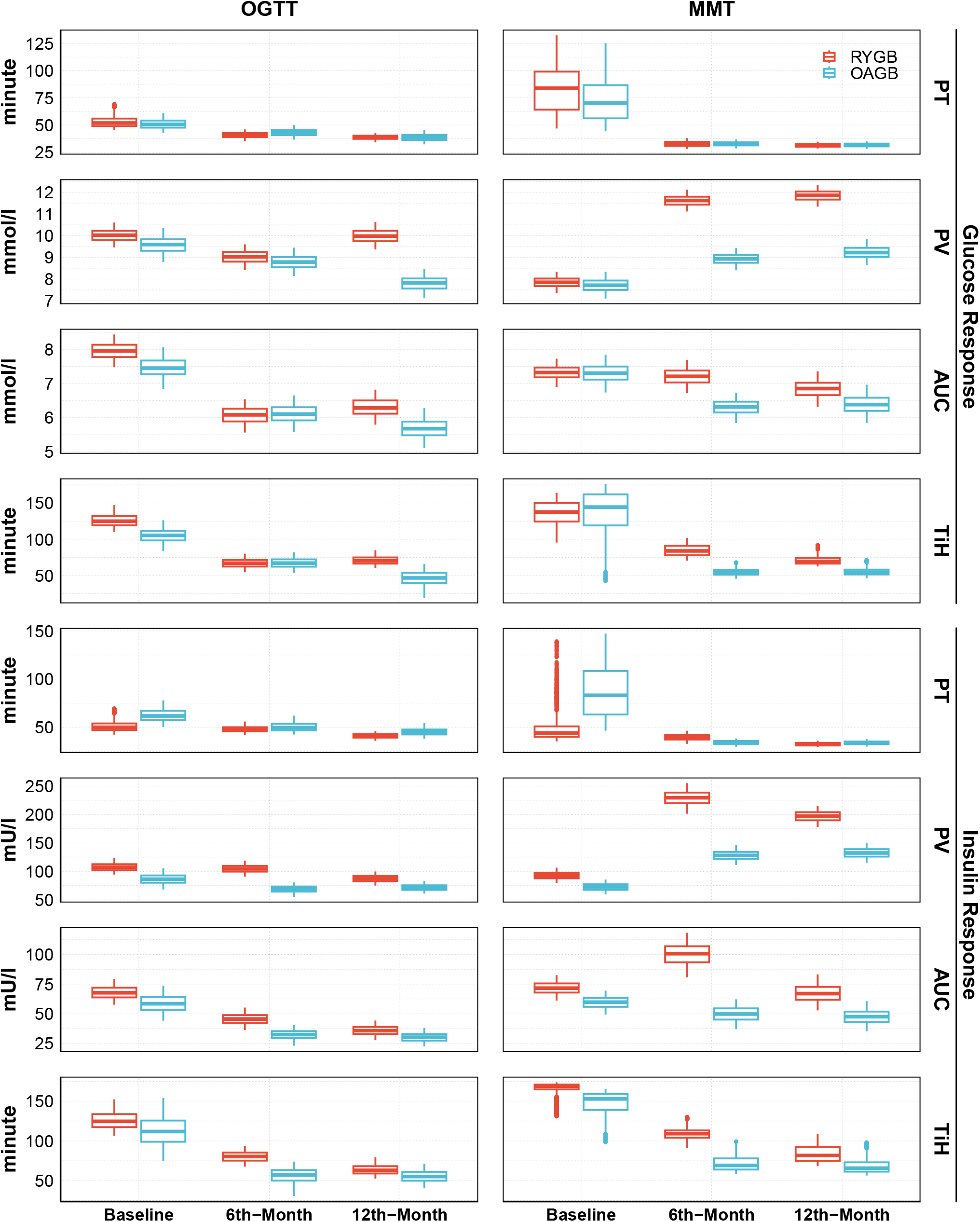
The box-plot of each metric for operation comparison. See Appendix F for the 95 % CIs and *p*-values.

In MMT, the RYGB group had similar PV of glucose but slightly higher PV of insulin than the OAGB group at baseline (Table F.2). During weight loss, the enhanced glucose and insulin responses in RYGB vs. OAGB were pronounced, with significantly higher PVs (Table F.2) and AUCs (Table F.3) of both glucose and insulin at 6th and 12th months. PTs of glucose and insulin were similar between the groups at all visits (Table F.1). Interestingly, the RYGB group experienced more TiH for glucose at the 6th month (83 min vs. 53 min) and a trend towards it at the 12th month (69 min vs. 54 min, p=0.054) compared with the OAGB group, despite similar baseline values between the groups (Table F.4). This may be partly due to the higher PV of glucose observed in RYGB. Over the 12 months, the reduction in TiH for glucose was 64 minutes for RYGB and 82 minutes for OAGB.

Overall, RYGB surgery induced a more pronounced glucose and insulin response in both tests, particularly in MMT, compared with OAGB, suggesting clinical differences between the procedures that can be taken into account in operation selection. Additionally, the pronounced PV and AUC in the RYGB group may drive the differences between MMT and OGTT in the postprandial response after surgery.

### 3.4 Higher postprandial glucose but not insulin response in men vs. women

Next, to account for other metabolic subgroups included in our cohort, we modeled women (n=30) and men (n=14) as different groups. Figure E.1 and Figure E.2 show population comparison for OGTT and MMT tests, respectively. In OGTT, men exhibited a higher AUC of insulin and higher PVs of glucose and insulin compared to women at baseline (Table F.3). Men had consistently higher postprandial PV of glucose response at all visits (Table F.2). At the 12th month, men had a higher AUC of glucose (Table F.3), later PTs for both glucose and insulin (Table F.1), and longer TiH for glucose with a smaller reduction in hyperglycemic time compared with women (31 min vs. 62 min) from baseline to 12th month (Table F.4).

In MMT, men and women showed similar baseline characteristics (Figure E.2). Men exhibited higher AUCs of glucose and insulin at the 6th month (Table F.3) along with higher PV of glucose at the 6th and 12th months (Table F.2) compared with women. At the 6th month, the PTs of glucose and insulin occurred later with men (Table F.1), and men also had longer TiH for glucose. However, the sexes had similar improvement in TiH during the weight loss (Table F.4).

### 3.5 Higher postprandial glucose but lower insulin response in T2DM vs. non-T2DM

We additionally evaluated the postprandial glucose and insulin responses between persons with T2DM (n=18) separately from those without (n=26) (Appendix D). As expected in OGTT, at all visits, persons with T2DM had higher PV and AUC, later PT and longer TiH for glucose, and later PT and lower PV for insulin (Appendix F). In MMT, PV and AUC of glucose were higher, and TiH for glucose was longer with T2DM at all visits. PV of insulin was lower than in persons without T2DM at the 12th month (Appendix F). Both groups improved in their glucose and insulin responses during weight loss.

### 3.6 Metabolic features in all participants improve during weight loss

At baseline, both RYGB and OAGB groups had similar metabolic features. Women had a higher fat percentage than men, and persons with T2DM had higher Hba1c and fasting plasma glucose than persons without T2DM. During the weight loss, body weight, fat kilograms, fat percentage, fasting glucose, insulin, and lipid levels improved significantly in all groups (Table G.1). A slightly more prominent decrease in Hba1c in favor of OAGB vs. RYGB was observed, and HDL-cholesterol, fat kilograms, and fat percentage improved better in men vs. women. In persons with T2DM weight, Hba1c and fasting glucose improved more than in those without T2DM. (Table G.1).

### 3.7 HMOGP model outperforms the conventional trapezoidal data analysis

We additionally analysed the data by conventional methods of estimating PT, PV and calculating the AUC and TiH during the test with linear trapezoid method. The comparison with HMOGP showed that HMOGP consistently predicted AUC and TiH with improved uncertainty compared to the conventional data analysis. Additionally, HMOGP was able to give realistic PT and PV estimations compared to the linear trapezoid method, as these parameters only relied on the timing of the observations in fundamental statistical analysis. Appendix H shows the comparison of HMOGP predictions with trapezoidal data analysis.

## 4 Discussion

We applied hierarchical multi-output Gaussian process (HMOGP) regression to analyze glucose and insulin dynamics in OGTT and MMT, identifying distinct metabolic responses based on bariatric surgery type and sex. Our findings reveal sharper and earlier glucose-insulin peaks with faster restoration to baseline in both OGTT and MMT post-surgery, along with more pronounced postprandial glucose-insulin responses in MMT than OGTT and in RYGB compared to OAGB. Our findings underscore RYGB’s role in enhanced glycemic control, while OAGB could be preferred in milder metabolic disturbances or when hypoglycemia may be a risk. RYGB’s sharper glucose peaks may have vascular implications, favoring OAGB for participants with cardiovascular disease. Additionally, we observed sex-based metabolic differences with men exhibiting a more unhealthy glucose-insulin profile throughout weight loss and distinctions between individuals with and without T2DM. We thus present that the HMOGP approach can be used to model postprandial glucose-insulin responses in detail in different groups of participants for personalized treatment strategies, allowing new means of understanding and improving individual glycemic responses in metabolic conditions.

Our study proposes the hierarchical multi-output Gaussian process (HMOGP) regression machine learning model as a suitable tool for modeling postprandial glucose-insulin data in detail for both OGTT and MMT. Previous studies have used the standard Gaussian process for modeling OGTT responses in participants with and without cystic fibrosis (Garrish et al., 2023), but HMOGP has not been applied to OGTT or MMT in relation to obesity or weight loss. Traditional linear regression models or simple curve-fitting approaches assume independence between glucose and insulin and fail to capture individual variability. Non-hierarchical Gaussian processes do not handle individual variations as effectively as the hierarchical structure in HMOGP (Alvarez and Lawrence, 2011; Williams and Rasmussen, 2006). Additionally, fixed-effect models like ANOVA, or simpler mechanistic models, cannot accommodate individual-to-population hierarchy and multi-output dependencies. In conventional linear trapezoid calculations, the outcomes rely strongly on the exact measurement time points. Our HMOGP method will allow a more granular understanding of how a patient compares with the population in insulin and glucose responses after surgery. The additional advantages are suitability for the correlated structure of glucose and insulin dynamics, for datasets with noise or missing values (typical in OGTT and MMT), improved accuracy of the prediction, and the ability to handle complex, multivariate data.

Both OGTT and MMT assess the pancreatic beta-cell response, which includes activation of the gut-related insulin-stimulating hormones. While OGTT is the standard for evaluating glucose intolerance and T2DM, MMT provides a more comprehensive physiological stimulus, as glucose combined with protein and fats enhance insulin secretion (Cersosimo et al., 2014). The duration of the postprandial glucose and insulin peaks were significantly elevated and shortened after both metabolic tests, confirming enhanced insulin stimulus and improved glucose clearance from the circulation post-operatively. Our results align with previous studies reporting that RYGB leads to a rapid increase in both postprandial blood glucose and insulin levels (Ferrannini and Mingrone, 2009). These metabolic benefits likely stem from gut reconfiguration post-surgery, including altered nutrient absorption and enhanced stimulation of L-cells, leading to a 10-fold increase in insulin-stimulating GLP-1 secretion (Cummings et al., 2007), and an increase in other gut hormones (Sandoval and Patti, 2023). Gut mucosa hyperplasia post-operation promotes glucose absorption (Cavin et al., 2016) while improved pancreatic islet coordination may enhance insulin release (Akalestou et al., 2021), collectively accelerating insulin secretion and glucose clearance. The enhanced post-surgical glucose-insulin response, earlier peak timing, and faster glucose normalization offer significant benefits, as hyperglycemia is linked to vascular dysfunction, cardiovascular risk, oxidative stress, inflammation, and cognitive decline (Di Giuseppe et al., 2023). Interestingly, earlier timing of the glucose-insulin peaks in OGTT also seems to indicate the effectiveness of the antidiabetic drugs (Tran et al., 2018) and reflect improved insulin sensitivity and secretion in T2DM (Wang et al., 2018b), while delayed glucose peaks reflect pancreatic beta-cell dysfunction (Tran et al., 2018). In addition, increasing OGTT curve complexity is associated with better glucose tolerance (Tura et al., 2011). However, sharp glycemic peaks after OGTT have also been linked to atherosclerosis (Temelkova-Kurktschiev et al., 2000) and higher pulse pressure (Anan et al., 2008), suggesting potential risks in certain metabolic conditions. The metabolic impact of such peaks in MMT remains unexplored. The metabolic and predictive value of the sharp and early glucose-insulin peaks will be better understood in the future.

In this study, RYGB resulted in a higher peak glucose-insulin response, larger AUC, and prolonged glucose levels above 7.0 mmol/l compared to OAGB, though both procedures showed similar peak timing and return to baseline. This difference was particularly evident in MMT post-surgery. While earlier non-randomized studies suggested a superior glucose profile after OAGB due to differences in bypassed intestine length (Wang et al., 2018a), randomized controlled trials (RCTs) with comparable anatomy have found RYGB and OAGB to be metabolically equivalent at 1- and 2-year follow-ups (Heinonen et al., 2023; Robert et al., 2019). The RCT on the same data as in this study reports similar improvements in fasting glucose, Hba1c, and postprandial glucose and insulin AUCs in OGTT (Heinonen et al., 2023) as well as comparable glycemic responses to carbohydrate intake measured by continuous glucose monitoring (Ashrafi et al., 2021). Our findings suggest that the more pronounced glucose-insulin peak in RYGB stems from its anatomical differences, where nutrients reach key gut areas more rapidly, leading to faster glucose absorption and a sharper insulin response compared to OAGB. This accelerated glycemic response may enhance early glucose control. In contrast, OAGB’s more gradual glucose absorption presents a smoother curve and may thus lower post-bariatric hypoglycemia risk, making it a preferable option for participants with preserved beta-cell function or milder metabolic disturbances. Although higher glycemic peaks in RYGB are linked to vascular risks like atherosclerosis and hypertension (Temelkova-Kurktschiev et al., 2000; Anan et al., 2008), studies indicate better hypertension improvement and reduced antihypertensive medication use post-RYGB (Heinonen et al., 2023). Given these dynamics, OAGB could, however, be a safer choice for individuals with a history of cardiovascular disease. Nonetheless, bariatric surgery remains a powerful metabolic intervention, providing substantial glycemic benefits regardless of the procedure (Sinclair et al., 2018).

Men exhibited higher peak glucose in OGTT compared to women, higher peak glucose and insulin, and longer TiH post-operatively in both OGTT and MMT, suggesting a more unhealthy profile in obesity and after weight loss. Typically, women have lower fasting plasma glucose, higher 2-hour glucose, and greater increases in 2-hour glucose following OGTT than men, though HbA1c levels are reported comparable between sexes (Sicree et al., 2008). Insulin sensitivity is also generally better in women (Nuutila et al., 1995). Research on obesity is sparse, with older studies indicating that men have higher peak glucose at 1 hour during OGTT, but women surpass them at 2-3 hours (Young et al., 1979). Recent data shows that bariatric surgery leads to similar T2DM resolution between sexes, though only in men, AUC of insulin during OGTT predicts weight loss (Risi et al., 2022; Shu et al., 2024). Previous studies have not compared OGTT and MMT responses before and after bariatric surgery and between sexes. We confirm a blunted glucose-insulin response in both sexes in obesity, an unhealthier glucose profile for men throughout the weight loss, and sex-related advantages with sharper and quicker glucose-insulin responses post-operatively in favor of women.

Bariatric surgery results in excellent rates of T2DM remission (Courcoulas et al., 2024), but its effects on postprandial glycemia between persons with or without T2DM are not yet well characterized. Some studies suggest less weight loss in persons with T2DM (Rebelos et al., 2023) but do not assess postprandial glycemic response. In our study, individuals with T2DM lost more weight and showed better metabolic improvements but had higher glucose, lower insulin responses, and a flatter curve at all visits compared to non-diabetic individuals. This altered response reflects significant changes in glucose and insulin dynamics in T2DM, warranting more individualized characterization of postprandial glycemic responses.

The strengths of this study include the standardized OGTT and MMT measurements on the same individuals undergoing bariatric surgery, with follow-up extending to 6th and 12th months. The used cohort and its subgroups are representative of a general population undergoing bariatric surgery (Heinonen et al., 2023; Saarinen et al., 2019) making the results generalizable. The comparisons between RYGB and OAGB are from a randomized controlled trial designed to compare these procedures, using identical bypass limb lengths and equal stratification of participants by sex, age, and T2DM status. These factors enhance the reliability of comparisons. Additionally, the MMT’s extended follow-up period of 360 minutes is a unique strength of this study. Analyzing responses across time and in subgroups further improves relevance by identifying procedure-specific, sex-related, and diabetes-associated patterns. While a larger, more diverse cohort would allow finer subgroup analyses, this study still provides valuable insights. Future research with an expanded dataset across different ethnicities could validate these findings. It is also worth noting that OGTT and MMT were performed on consecutive days, which may introduce slight day-to-day variation.

## 5 Conclusions

Utilizing Hierarchical Multi-Output Gaussian Process (HMOGP) regression modeling to analyze postprandial OGTT and MMT responses presents an advanced approach to studying postprandial glucose-insulin responses in obesity and weight loss. The employed HMOGP regression is capable of estimating individual-level effects — the personalized treatment response — in addition to the effects at the population level while accounting for and correcting for measurement errors and thus improving accuracy in complex multivariate data. This method not only unveils differences between metabolic cohorts after bariatric surgery but also a differential response between two operation types and sexes. This can lead to new knowledge in choosing the right treatments for participants and a more accurate modeling method for assessing OGTT and MMT responses in the clinic. Our study suggests that based on postprandial glucose dynamics, RYGB may be favorable over OAGB in hyperglycemic patients without risk for hypoglycemia or vascular diseases, where OAGB could be preferred. Thus, HMOGP regression offers a versatile tool for investigating and predicting glycemic responses in metabolic conditions, paving the way for personalized interventions in metabolic health.

## Supporting information

Supplemental Table G.1 - detailed

## Data Availability

Individual participant data are not directly available but may be available after study close upon reasonable request from the authors. According to the General Data Protection Regulation of the European Union (679/2016), the principles of data protection should apply to any information concerning an identified or identifiable natural person and that personal data which have undergone pseudonymization, which could be attributed to a natural person by the use of additional information should be considered to be information on an identifiable natural person. Thus, according to the GDPR, all pseudonymized data are considered personal data and cannot be published openly. Therefore, we are bound to the law and to the strict hospital policies, and are unable to share the data directly. However, the institutional (Helsinki and Uusimaa Hospital District) contact details for potential future data requests are as follows: https://huspalvelu.microsoftcrmportals.com/fi-FI/

## Acknowledgments

We thank the participants for their invaluable time and effort. We also thank the study nurses, dieticians, and laboratory personnel for their help.

## A Modeling Details

We normalized the data in both time and value domains before the training, separately for insulin and glucose values. We used a fixed lengthscale parameters for all GPs, which is set to be 120*/σ*_time_, where *σ*_time_ is the standard deviation of the sample times. We used the Matern52 kernel for all GPs. The main difference between individual-level GPs and the population-level GPs was the prior value of the kernel variance. We used *InverseGamma*(1, 1) distribution for the population level GPs, while for the individual level GPs, we used a stronger *InverseGamma*(1, 0.1) distribution. This setup allowed the model to learn the shared behaviors with the population-level GPs while the individual-level GPs learned individual differences. For measurement error *σ*, we used *InverseGamma*(1, 0.01) as a prior distribution. For ***µ***^population^, we used 𝒩(0, 1) as a prior distribution, and for 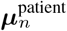 we used 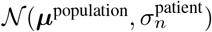 as a prior distribution, where 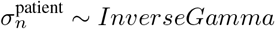 (1, 0.5). The rest of the details and full Stan code are available at https://github.com/onurpoyraz/HMOGP.

## B Individual Responses

**Figure B.1.**
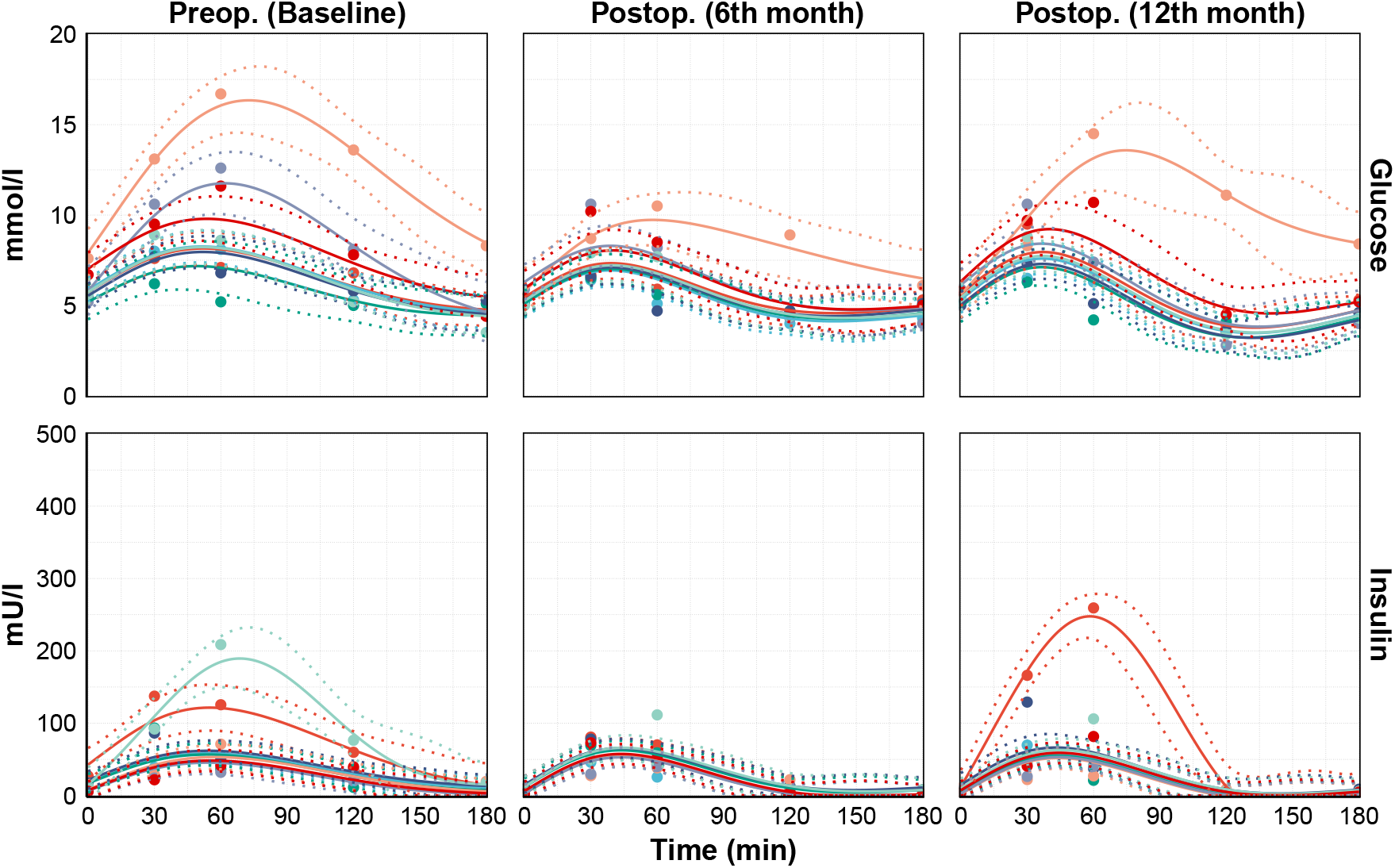
Response of the glucose and insulin levels and predicted model fit for different individuals with applied OGTT test.

**Figure B.2.**
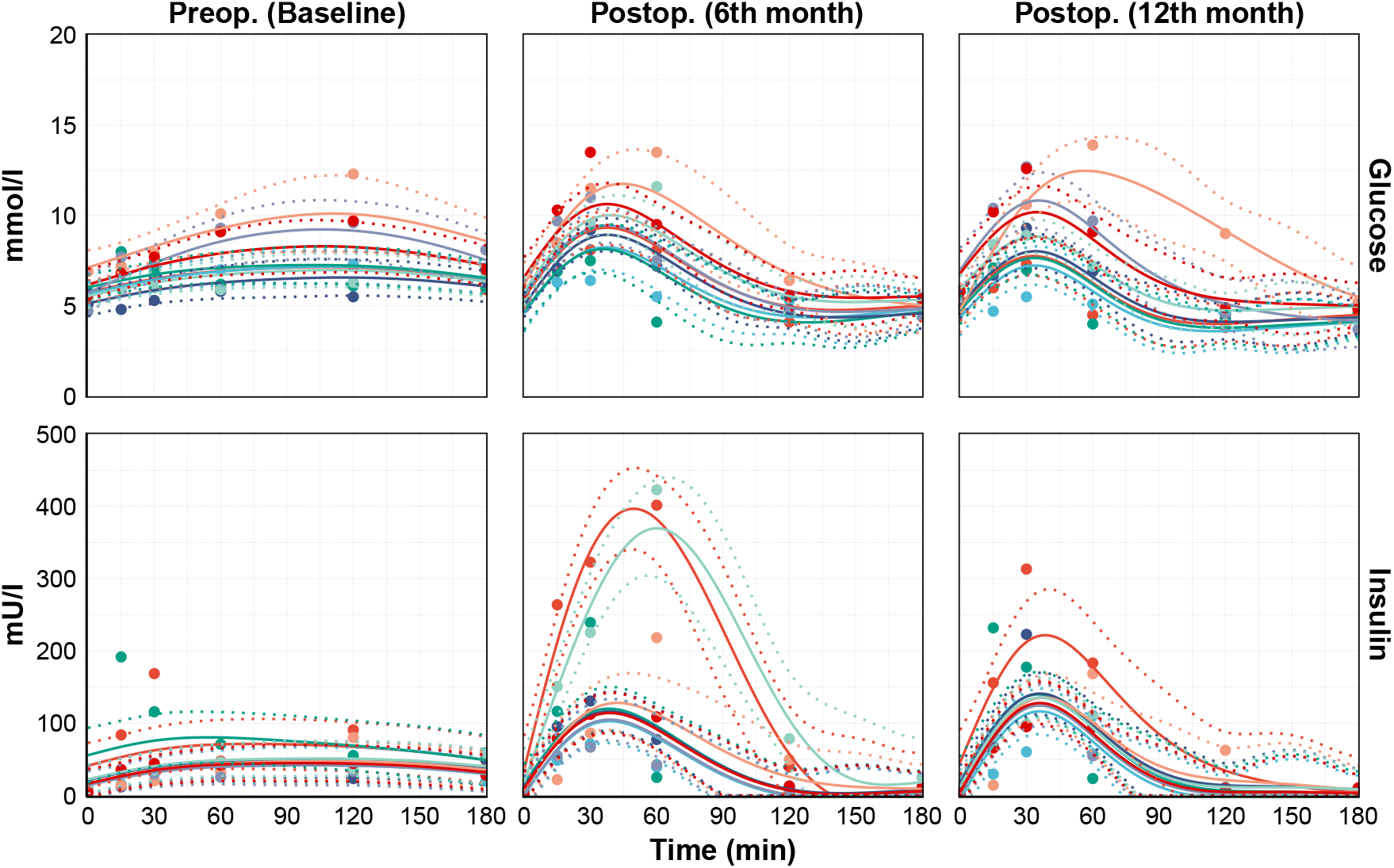
Response of the glucose and insulin levels and predicted model fit for different individuals with applied MMT test.

## C MMT Model Fitting

MMT includes additional data which sampled at 15th, 240th, and 360th minutes compared to OGTT. So, for an healthy comparison we trained the model on MMT using the same datapoints exist on the OGTT (partial data). Figure C.1 compares the differences between the model fits with the full data and partial data on MMT.

**Figure C.1.**
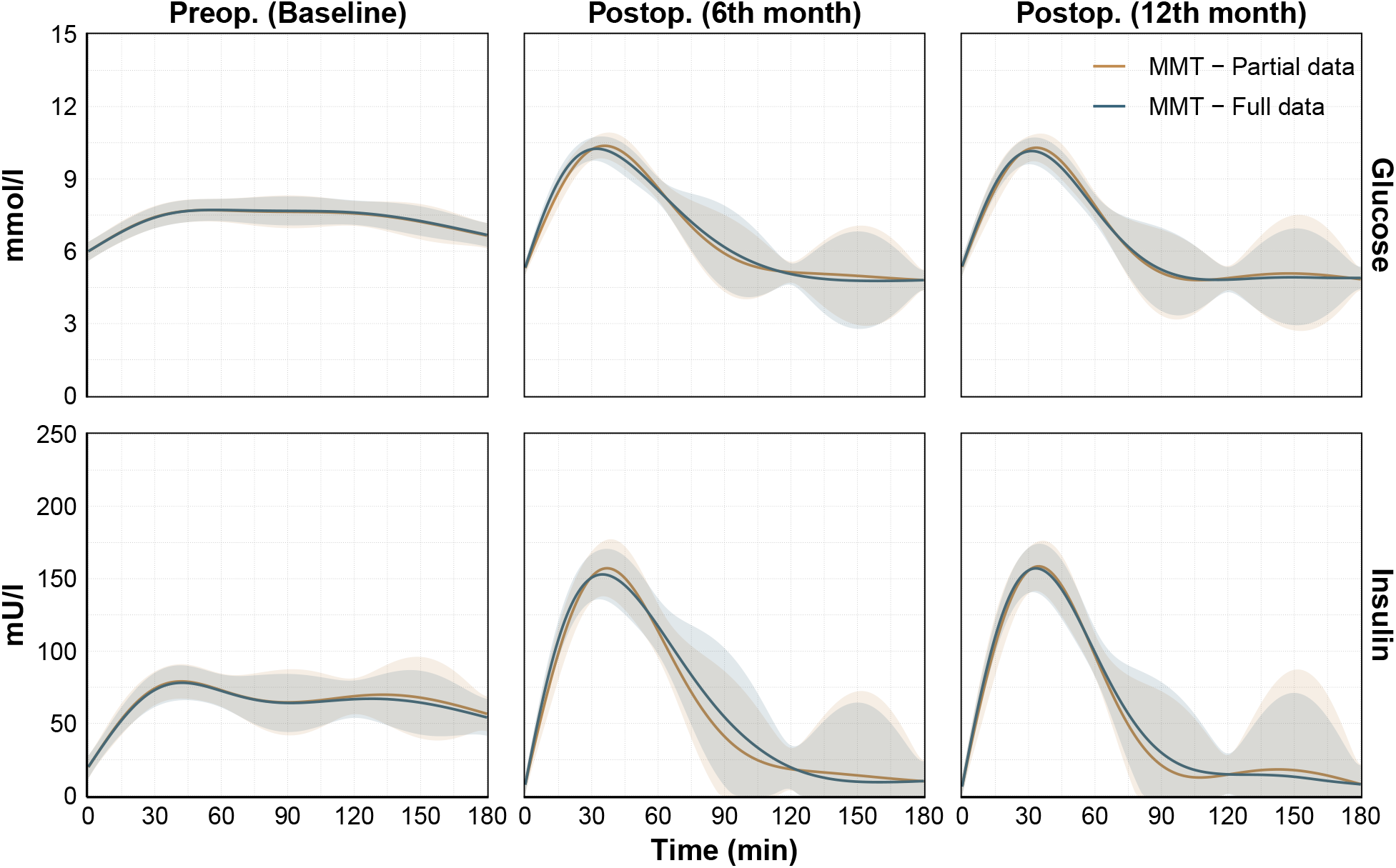
Difference of model predictions with partial data vs. full data on the MMT. Shaded areas represent 95% confidence intervals (CI).

## D Comparison by Diabetes

**Figure D.1.**
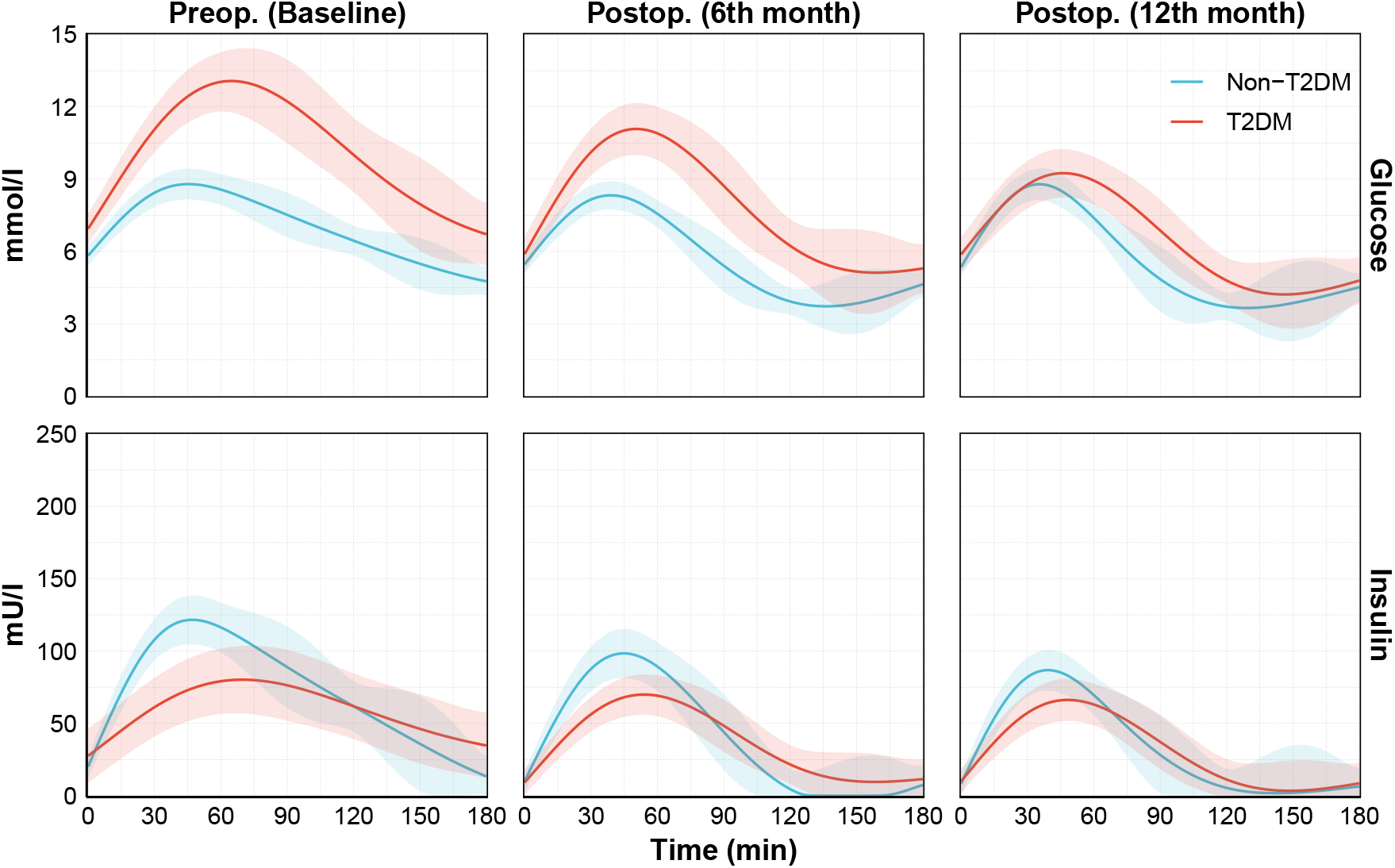
The comparison of model predicted average glucose and insulin response of the Non-T2DM and T2DM participants in OGTT test. Shaded areas represent 95% confidence intervals (CI).

**Figure D.2.**
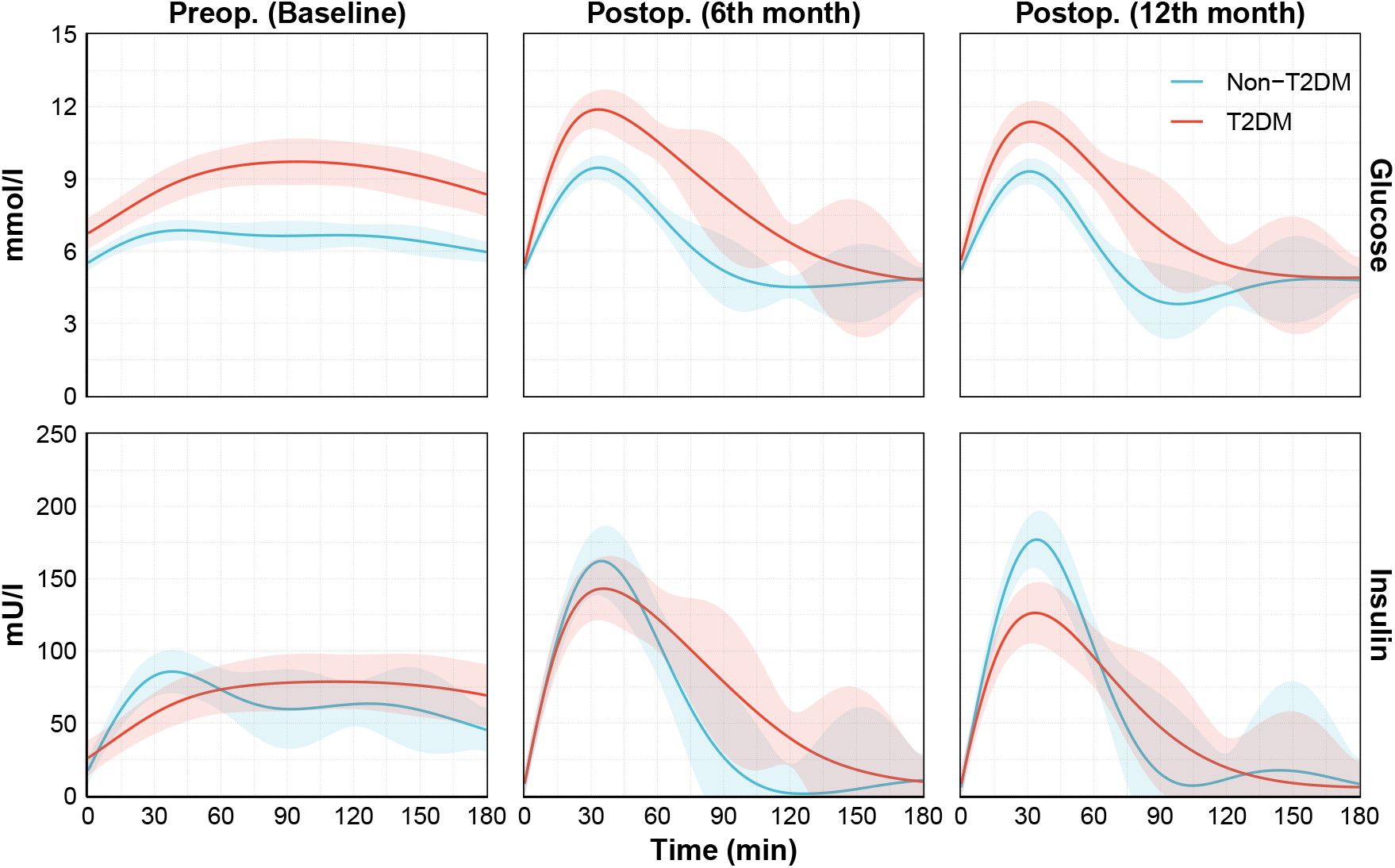
The comparison of model predicted average glucose and insulin response of the Non-T2DM and T2DM participants in MMT test. Shaded areas represent 95% confidence intervals (CI).

**Figure D.3.**
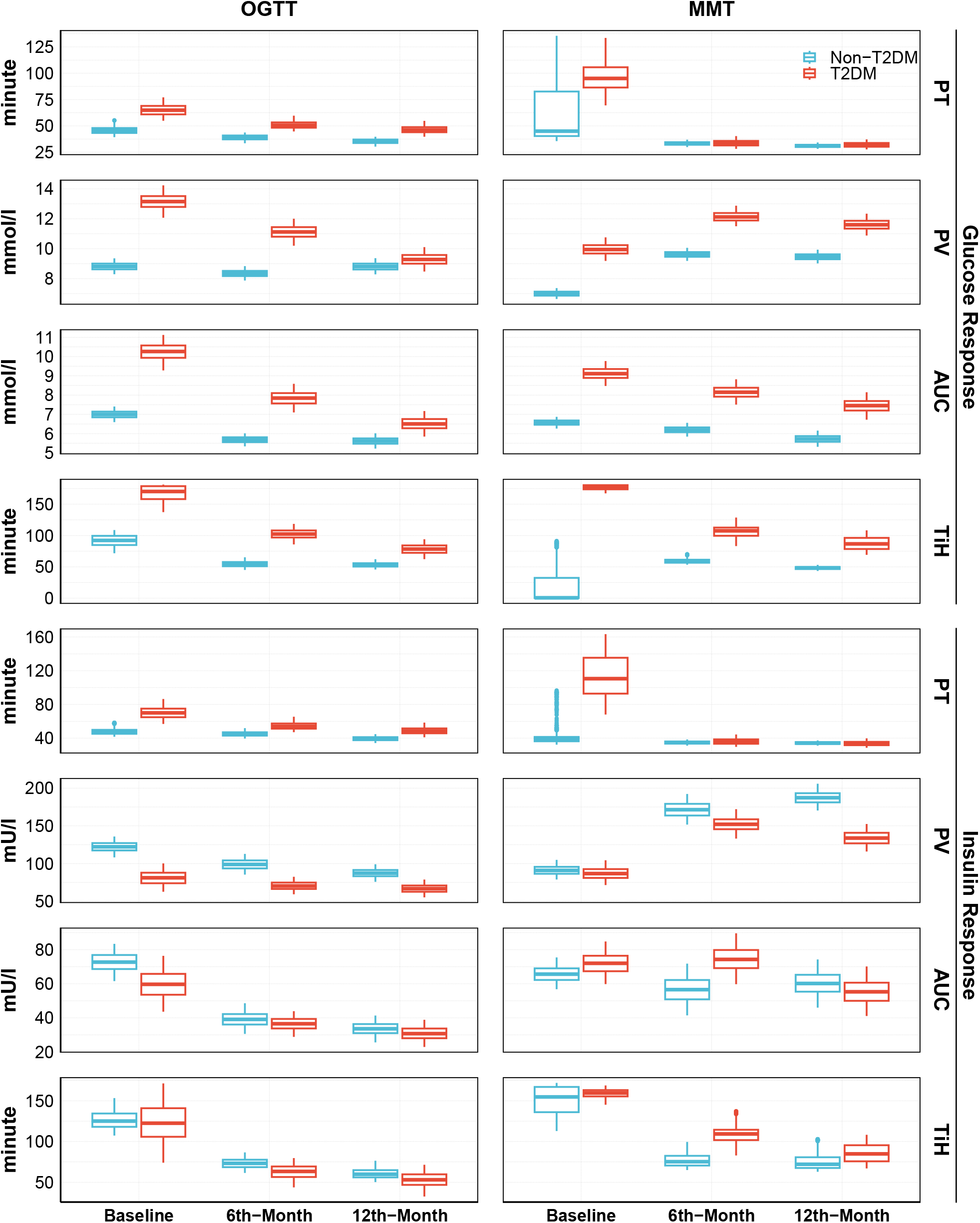
The box-plot of each metric for T2DM comparison. See Appendix F for the 95% CIs and *p*-values.

## E Comparison by Sex

**Figure E.1.**
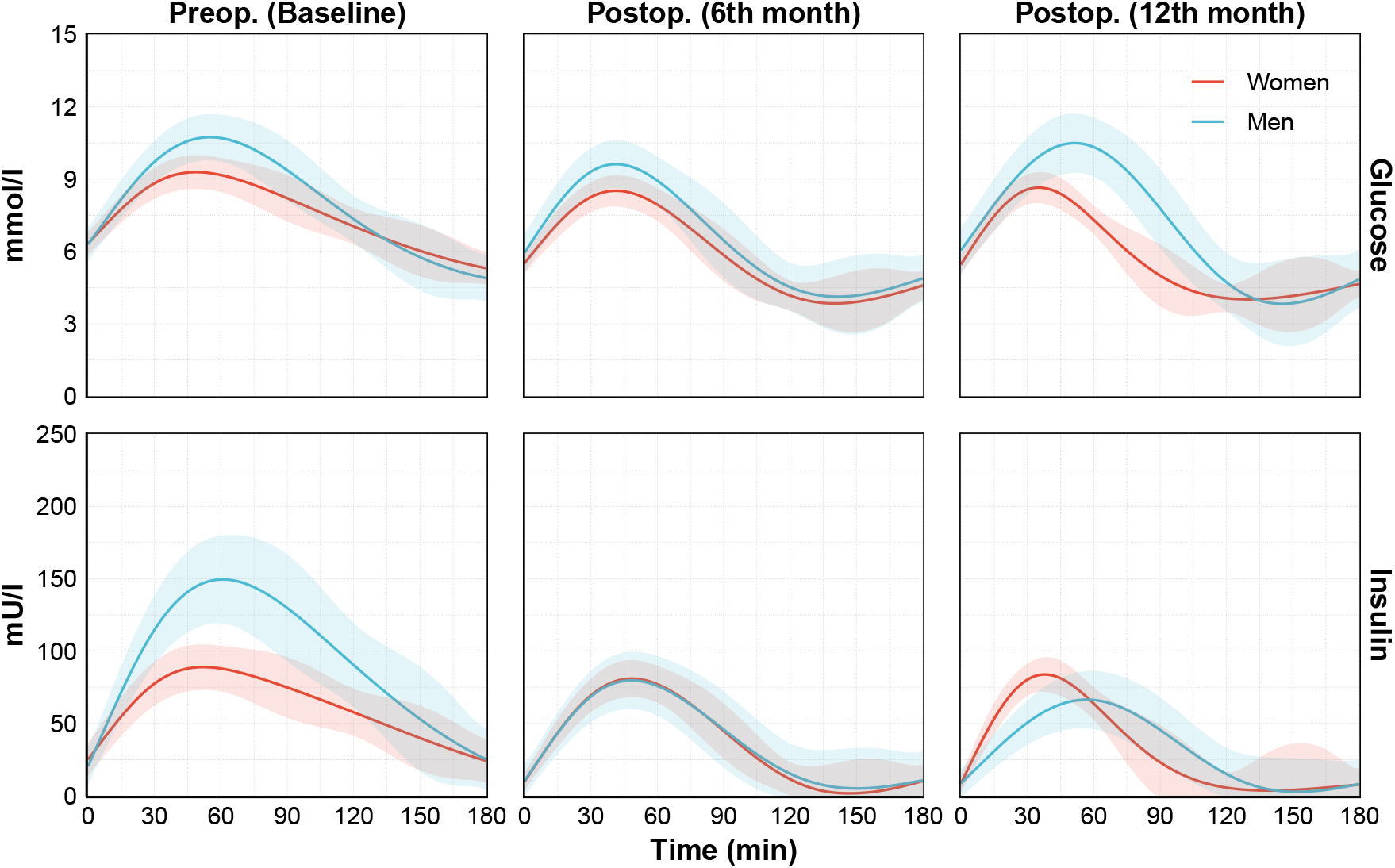
The comparison of model predicted average glucose and insulin response of the women and men participants in OGTT test. Shaded areas represent 95% confidence intervals (CI).

**Figure E.2.**
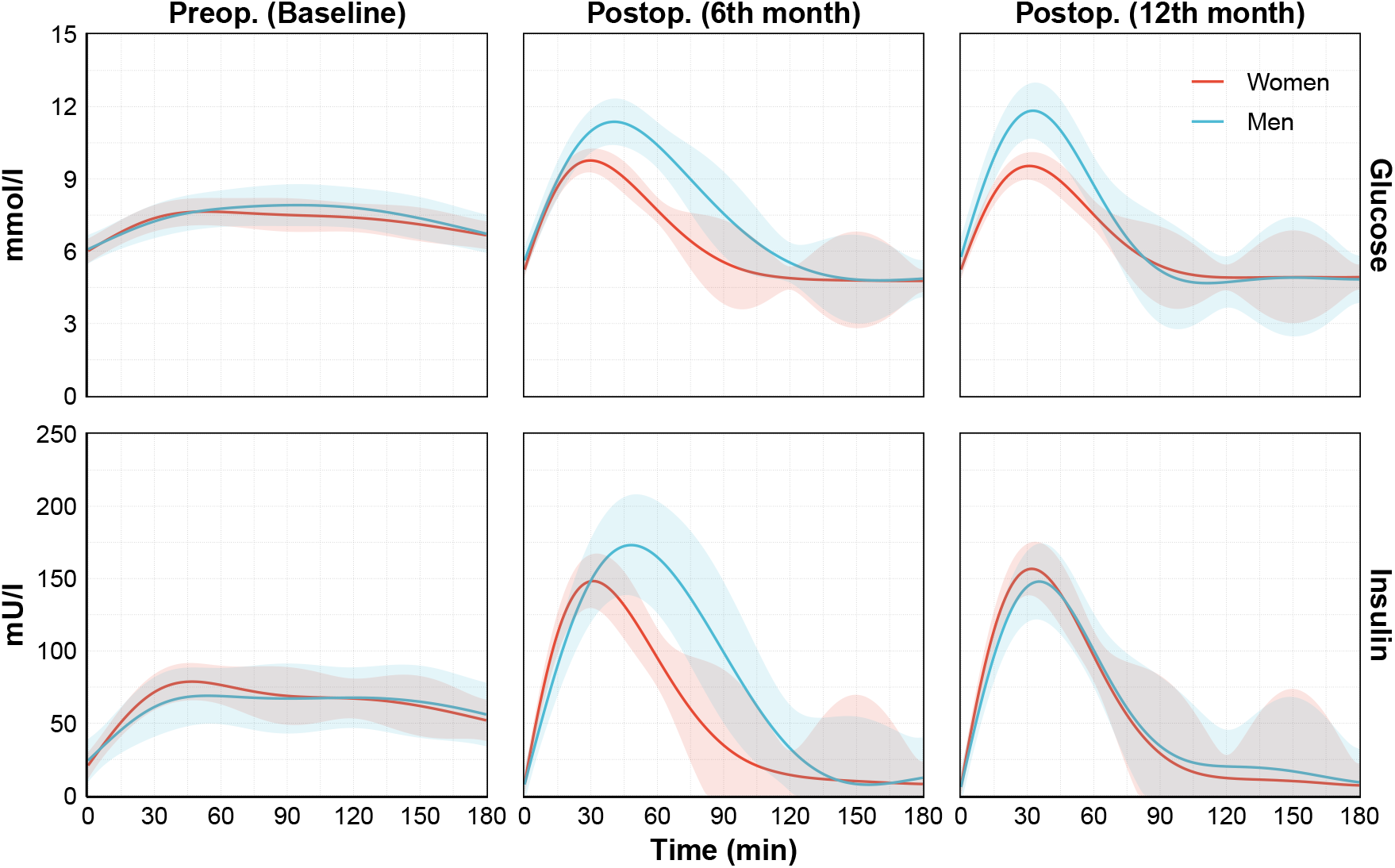
The comparison of model predicted average glucose and insulin response of the women and men participants in MMT test.

**Figure E.3.**
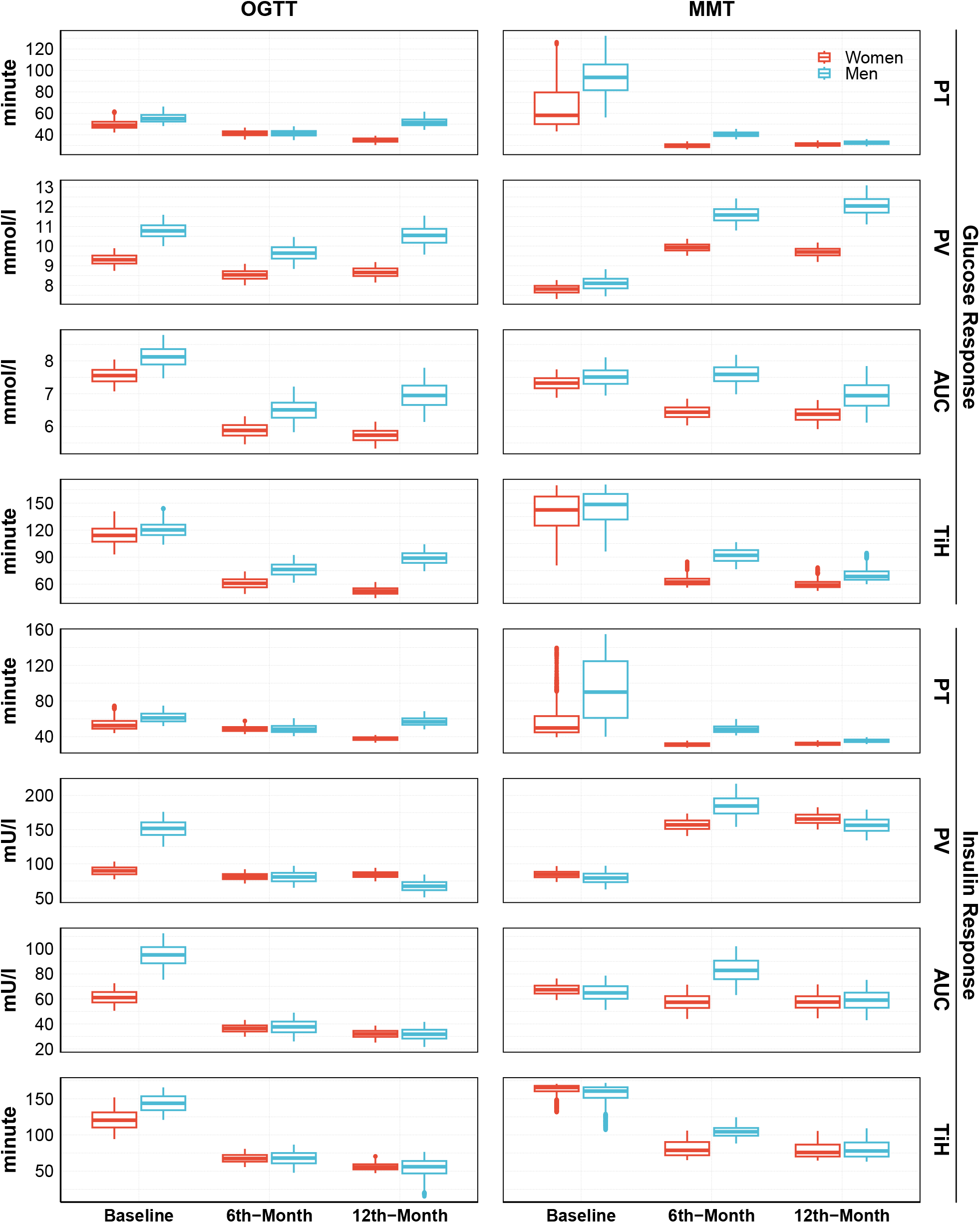
The box-plot of each metric for sex comparison. See Appendix F for the 95% CIs and *p*-values.

## F Evaluation of Metrics

**Table F.1.**
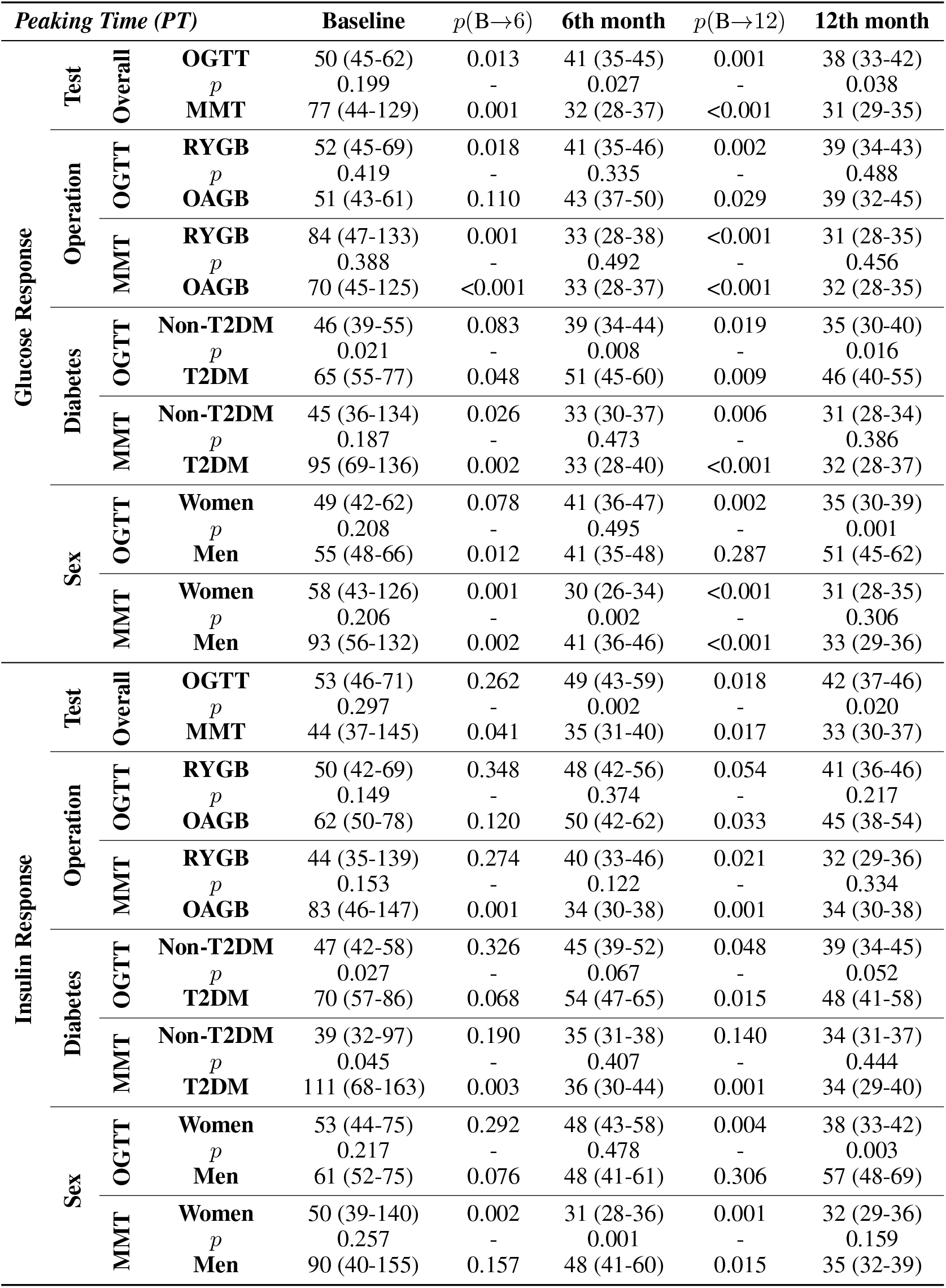
Peaking time (PT, minute) of the glucose and insulin responses with 95% confidence intervals and *p*-values. *p*-values (except *p*(B *→* 12)) shows the statistical significance of the difference between adjacent cells. *p*(B *→* 12) represents the statistical significance of the difference between Baseline and 12th month values.

**Table F.2.**
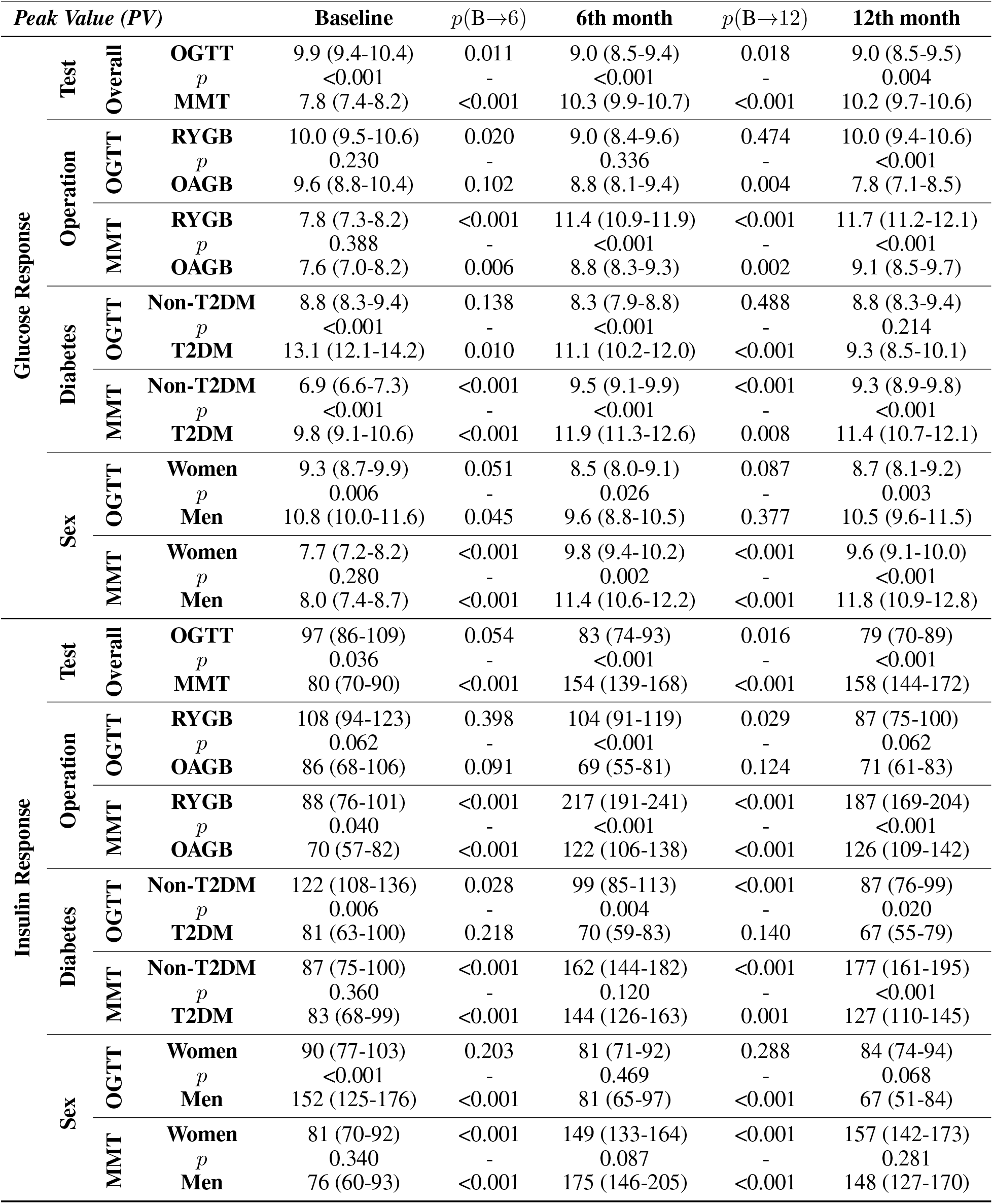
Peak value (PV, mmol/l for glucose, mU/l for insulin) of the glucose and insulin responses with 95% confidence intervals and *p*-values. *p*-values (except *p*(B*→*12)) shows the statistical significance of the difference between adjacent cells. *p*(B*→*12) represents the statistical significance of the difference between Baseline and 12th month values.

**Table F.3.**
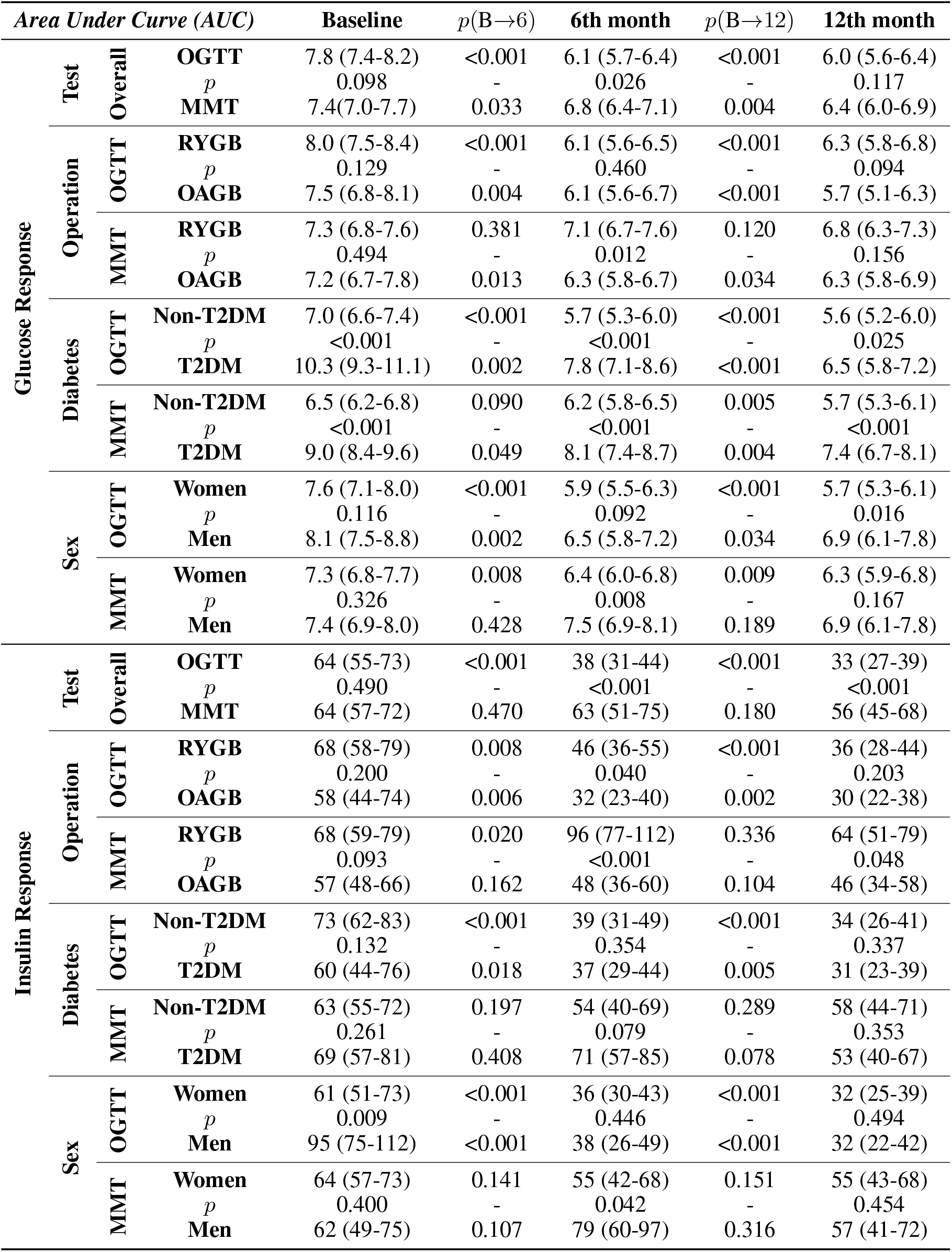
Time-normalized Area Under Curve (AUC), mmol/l for glucose, mU/l for insulin) of the glucose and insulin responses with 95% confidence intervals and *p*-values. *p*-values (except *p*(B*→*12)) shows the statistical significance of the difference between adjacent cells. *p*(B*→*12) represents the statistical significance of the difference between Baseline and 12th month values.

**Table F.4.**
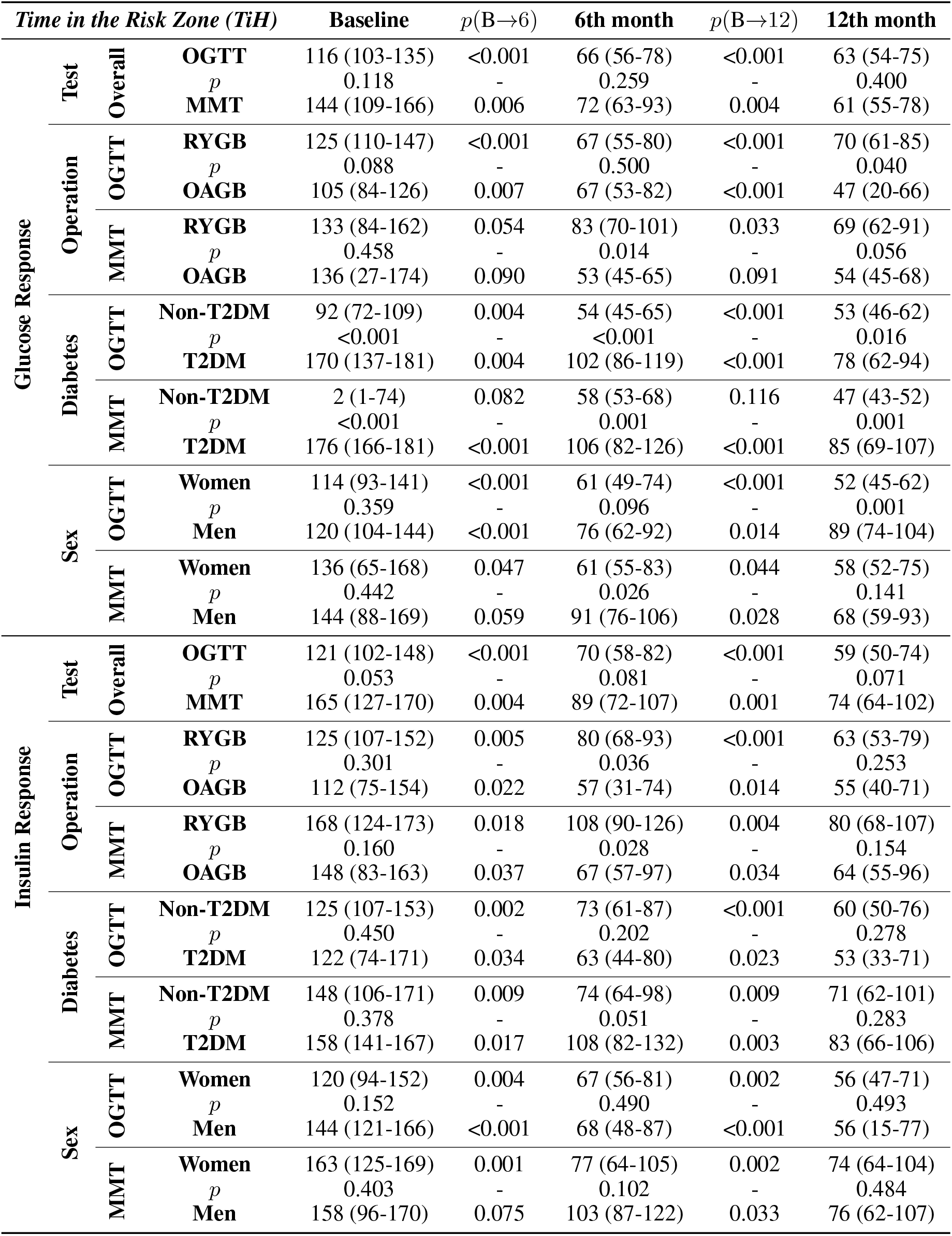
Time in the Risk Zone (TiH, minute), where hyperglycemia and hyperinsulinemia are defined as glucose > 7.0mmol/l and insulin > 50mU/l, respectively, with 95 % confidence intervals and *p*-values. *p*-values (except *p*(B *→* 12)) shows the statistical significance of the difference between adjacent cells. *p*(B *→* 12) represents the statistical significance of the difference between Baseline and 12th month values.

## G Data Statistics

**Table G.1.** Metabolic features of the participants by different operations, sexes, and diabetes over each visit. **B** represents baseline visit, **6th** corresponds to 6th month visit, and **12th** shows 12th month visit.

| *Metabolic Features* |  | **Overall** | **OAGB** | **RYGB** | **Male** | **Female** | **T2DM** | **Non-T2DM** |
| --- | --- | --- | --- | --- | --- | --- | --- | --- |
| **Age (years)** | **B** | 46.0 ± 7.0 | 46.6 ± 7.4 | 45.4 ± 6.6 | 50.7 ± 7.4 | 43.9 ± 5.8 | 44.3 ± 6.3 | 47.3 ± 7.3 |
|  | **6th** | 46.5 ± 7.0 | 47.1 ± 7.4 | 45.9 ± 6.6 | 51.2 ± 7.4 | 44.4 ± 5.8 | 44.8 ± 6.3 | 47.8 ± 7.3 |
|  | **12th** | 47.0 ± 7.0 | 47.6 ± 7.4 | 46.4 ± 6.6 | 51.7 ± 7.4 | 44.9 ± 5.8 | 45.3 ± 6.3 | 48.3 ± 7.3 |
| **Weight (kg)** | **B** | 130 ± 19 | 127 ± 15 | 135 ± 23 | 138 ± 22 | 127 ± 17 | 132 ± 21 | 130 ± 18 |
|  | **6th** | 102 ± 16 | 99 ± 14 | 107 ± 19 | 109 ± 18 | 101 ± 16 | 101 ± 16 | 104 ± 17 |
|  | **12th** | 98 ± 16 | 94 ± 14 | 103 ± 18 | 106 ± 19 | 95 ± 15 | 95 ± 16 | 101 ± 17 |
| **Bmi (kg/m2)** | **B** | 45 ± 6 | 44 ± 6 | 46 ± 6 | 42 ± 5 | 46 ± 6 | 46 ± 6 | 44 ± 6 |
|  | **6th** | 35 ± 5 | 35 ± 6 | 36 ± 5 | 34 ± 5 | 37 ± 5 | 35 ± 5 | 36 ± 5 |
|  | **12th** | 34 ± 5 | 33 ± 6 | 35 ± 5 | 33 ± 5 | 35 ± 5 | 34 ± 6 | 35 ± 5 |
| **Hba1c (mmol/mol)** | **B** | 39 ± 10 | 38 ± 6 | 41 ± 13 | 38 ± 7 | 40 ± 11 | 47 ± 12 | 34 ± 4 |
|  | **6th** | 34 ± 5 | 34 ± 3 | 33 ± 6 | 34 ± 4 | 34 ± 5 | 37 ± 4 | 32 ± 4 |
|  | **12th** | 34 ± 7 | 34 ± 3 | 35 ± 10 | 35 ± 6 | 34 ± 7 | 38 ± 8 | 31 ± 4 |
| **Cholesterol (mmol/l)** | **B** | 4.5 ± 0.9 | 4.5 ± 1.0 | 4.6 ± 0.7 | 4.3 ± 1.3 | 4.6 ± 0.7 | 4.5 ± 1.1 | 4.5 ± 0.8 |
|  | **6th** | 3.8 ± 0.7 | 3.7 ± 0.7 | 3.9 ± 0.6 | 3.5 ± 0.8 | 3.9 ± 0.6 | 3.7 ± 0.8 | 3.9 ± 0.6 |
|  | **12th** | 3.9 ± 1.0 | 3.8 ± 0.7 | 4.3 ± 1.2 | 3.8 ± 1.1 | 4.1 ± 0.9 | 3.8 ± 0.9 | 4.1 ± 1.1 |
| **HDL Cholesterol (mmol/l)** | **B** | 1.2 ± 0.2 | 1.2 ± 0.2 | 1.2 ± 0.2 | 1.2 ± 0.2 | 1.3 ± 0.2 | 1.2 ± 0.3 | 1.2 ± 0.2 |
|  | **6th** | 1.2 ± 0.2 | 1.2 ± 0.2 | 1.3 ± 0.2 | 1.2 ± 0.2 | 1.2 ± 0.2 | 1.1 ± 0.2 | 1.3 ± 0.2 |
|  | **12th** | 1.4 ± 0.3 | 1.4 ± 0.2 | 1.4 ± 0.3 | 1.4 ± 0.3 | 1.4 ± 0.3 | 1.3 ± 0.3 | 1.5 ± 0.2 |
| **LDL Cholesterol (mmol/l)** | **B** | 2.9 ± 0.8 | 2.8 ± 0.9 | 3.0 ± 0.7 | 2.8 ± 1.1 | 2.9 ± 0.6 | 2.8 ± 0.9 | 2.9 ± 0.8 |
|  | **6th** | 2.3 ± 0.7 | 2.2 ± 0.7 | 2.4 ± 0.6 | 2.0 ± 0.7 | 2.4 ± 0.6 | 2.3 ± 0.7 | 2.3 ± 0.6 |
|  | **12th** | 2.3 ± 0.8 | 2.2 ± 0.8 | 2.3 ± 0.8 | 2.1 ± 1.1 | 2.3 ± 0.6 | 2.2 ± 0.8 | 2.3 ± 0.7 |
| **Triglyceride (mmol/l)** | **B** | 1.4 ± 0.7 | 1.4 ± 0.6 | 1.4 ± 0.8 | 1.3 ± 0.6 | 1.5 ± 0.7 | 1.7 ± 0.9 | 1.3 ± 0.5 |
|  | **6th** | 1.1 ± 0.4 | 1.1 ± 0.4 | 1.1 ± 0.3 | 0.9 ± 0.3 | 1.2 ± 0.3 | 1.2 ± 0.4 | 1.0 ± 0.3 |
|  | **12th** | 1.0 ± 0.3 | 0.9 ± 0.3 | 1.0 ± 0.3 | 0.9 ± 0.3 | 1.0 ± 0.3 | 1.1 ± 0.3 | 0.9 ± 0.3 |
| **Fasting Glucose (mmol/l)** | **B** | 6.1 ± 1.0 | 5.9 ± 0.9 | 6.3 ± 1.2 | 6.1 ± 0.9 | 6.1 ± 1.1 | 6.8 ± 1.1 | 5.6 ± 0.6 |
|  | **6th** | 5.4 ± 0.7 | 5.4 ± 0.6 | 5.4 ± 0.8 | 5.7 ± 0.8 | 5.3 ± 0.6 | 5.6 ± 1.0 | 5.3 ± 0.4 |
|  | **12th** | 5.4 ± 0.9 | 5.4 ± 0.8 | 5.5 ± 1.0 | 5.8 ± 1.2 | 5.3 ± 0.7 | 5.7 ± 1.2 | 5.2 ± 0.5 |
| **Fasting Insulin (mU/l)** | **B** | 20.3 ± 17.2 | 20.5 ± 14.7 | 20.1 ± 20.0 | 19.1 ± 14.5 | 20.9 ± 18.5 | 24.8 ± 23.6 | 17.2 ± 9.8 |
|  | **6th** | 8.2 ± 4.4 | 8.8 ± 4.6 | 7.7 ± 4.2 | 8.4 ± 4.6 | 8.2 ± 4.4 | 7.9 ± 3.9 | 8.5 ± 4.8 |
|  | **12th** | 6.6 ± 4.1 | 7.1 ± 4.6 | 6.2 ± 3.5 | 6.7 ± 3.7 | 6.7 ± 4.3 | 7.0 ± 4.1 | 6.4 ± 4.1 |
| **Fat mass (kg)** | **B** | 64 ± 13 | 62 ± 12 | 65 ± 15 | 61 ± 16 | 65 ± 12 | 64 ± 12 | 63 ± 14 |
|  | **6th** | 43 ± 12 | 41 ± 11 | 45 ± 12 | 39 ± 14 | 45 ± 10 | 39 ± 9 | 46 ± 13 |
|  | **12th** | 38 ± 12 | 36 ± 13 | 39 ± 12 | 31 ± 14 | 40 ± 11 | 35 ± 11 | 40 ± 13 |
| **Fat ratio (percent)** | **B** | 49 ± 6 | 49 ± 6 | 49 ± 7 | 44 ± 7 | 51 ± 4 | 49 ± 5 | 49 ± 7 |
|  | **6th** | 41 ± 7 | 41 ± 8 | 41 ± 7 | 35 ± 8 | 43 ± 5 | 39 ± 7 | 42 ± 8 |
|  | **12th** | 37 ± 9 | 37 ± 10 | 38 ± 9 | 30 ± 9 | 41 ± 6 | 36 ± 8 | 39 ± 9 |

## H Model Predictions vs. Data Distribution

**Table H.1.** The comparison of HMOGP predictions (model) with fundamental statistical analysis on the data distribution (data). The comparison shows that fundamental statistical analysis of the data distribution gives unreliable peaking time (PT) and peak value (PV) results as they only rely on the timing of the observations. On the other hand, HMOGP predicts consistent AUC and TiH estimations with improved uncertainty compared to the conventional data analysis.

| *Glucose Response* | | | Baseline | $p(B \rightarrow 6)$ | 6th month | $p(B \rightarrow 12)$ | 12th month | Unit |
| --- | --- | --- | --- | --- | --- | --- | --- | --- |
| Peaking Time<br>(PT) | Data<br>Statistics | OGTT | 60 (30-120) | 0.409 | 30 (30-60) | 0.417 | 30 (0-60) | minute |
| | | $p$ | 0.250 | - | 0.391 | - | 0.386 | |
|  |  | MMT | 60 (30-180) | 0.150 | 30 (30-60) | 0.179 | 30 (30-60) |  |
|  | HMOGP<br>Predictions | OGTT | 50 (45-62) | 0.013 | 41 (35-45) | 0.001 | 38 (33-42) |  |
| | | $p$ | 0.199 | - | 0.027 | - | 0.038 | |
|  |  | MMT | 77 (44-129) | 0.001 | 32 (28-37) | <0.001 | 31 (29-35) |  |
| Peak Value<br>(PV) | Data<br>Statistics | OGTT | 10.2 (6.2-16.5) | 0.364 | 9.8 (6.5-17.3) | 0.403 | 9.6 (6.3-21.7) | mmol/l |
| | | $p$ | 0.105 | - | 0.297 | - | 0.371 | |
|  |  | MMT | 8.0 (5.7-12.7) | 0.162 | 11.1 (7.4-16.5) | 0.205 | 10.7 (6.9-16.4) |  |
|  | HMOGP<br>Predictions | OGTT | 9.9 (9.4-10.4) | 0.011 | 9.0 (8.5-9.4) | 0.018 | 9.0 (8.5-9.5) |  |
| | | $p$ | <0.001 | - | <0.001 | - | 0.004 | |
|  |  | MMT | 7.8 (7.4-8.2) | <0.001 | 10.3 (9.9-10.7) | <0.001 | 10.2 (9.7-10.6) |  |
| Area Under Curve<br>(AUC) | Data<br>Statistics | OGTT | 7.4 (5.3-13.5) | 0.061 | 6.2 (4.9-11.9) | 0.222 | 6.1 (4.7-13.7) | mmol/l |
| | | $p$ | 0.368 | - | 0.312 | - | 0.371 | |
|  |  | MMT | 7.1 (5.6-11.1) | 0.300 | 6.9 (5.3-10.7) | 0.308 | 6.3 (4.9-11.3) |  |
|  | HMOGP<br>Predictions | OGTT | 7.8 (7.4-8.2) | <0.001 | 6.1 (5.7-6.4) | <0.001 | 6.0 (5.6-6.4) |  |
| | | $p$ | 0.098 | - | 0.026 | - | 0.117 | |
|  |  | MMT | 7.4 (7.0-7.7) | 0.033 | 6.8 (6.4-7.1) | 0.004 | 6.4 (6.0-6.9) |  |
| Time in the Risk Zone<br>(TiH) | Data<br>Statistics | OGTT | 104 (0-181) | 0.136 | 70 (0-179) | 0.236 | 60 (0-181) | minute |
| | | $p$ | 0.474 | - | 0.391 | - | 0.414 | |
|  |  | MMT | 120 (0-181) | 0.400 | 78 (10-172) | 0.397 | 62 (0-176) |  |
|  | HMOGP<br>Predictions | OGTT | 116 (103-135) | <0.001 | 66 (56-78) | <0.001 | 63 (54-75) |  |
| | | $p$ | 0.118 | - | 0.259 | - | 0.400 | |
|  |  | MMT | 144 (109-166) | 0.006 | 72 (63-93) | 0.004 | 61 (55-78) |  |

