## Supplemental Table G.1 - detailed for "Computational modeling enables individual assessment of postprandial glucose and insulin responses after bariatric surgery"

**Supplementary Metabolic table p-values**

p-values for difference between the groups at the investigated timepoint (Student's paired T-test)
p-values for difference between the groups from baseline to 6th or to 12th month with generalized Linear Mixed model.

Participants as random effects and adjusted for:
1) sex and T2DM status (OAGB vs. RYGB),
2) operation type and T2DM status (male vs. female),
3) operation type and sex (T2DM vs. nonT2DM).

| | | | | | | *between the groups* | | | | | *between the groups* | | | | | *between the groups* | | |
|---|---|---|---|---|---|---|---|---|---|---|---|---|---|---|---|---|---|---|
| **Age** | | **overall** | **SD** | **OAGB** | **SD** | **p-value** | **RYGB** | **SD** | **male** | **SD** | **p-value** | **female** | **SD** | **T2DM** | **SD** | **p-value** | **nonT2DM** | **SD** |
| **B** | **p-value at baseline** | 46 | ±7.0 | 46.6 | ±7.4 | 0.702 | 45.4 | ±6.6 | 50.7 | ±7.4 | 0.003 | 43.9 | ±5.8 | 44.3 | ±6.3 | 0.262 | 47.3 | ±7.3 |
| | **p-value (B→6)** | | | | | 0.301 | | | | | 0.348 | | | | | 0.995 | | |
| **6th** | **p-value at 6th month** | 46.5 | ±7.0 | 47.1 | ±7.4 | 0.568 | 45.9 | ±6.6 | 51.2 | ±7.4 | 0.004 | 44.4 | ±5.8 | 44.8 | ±6.3 | 0.178 | 47.8 | ±7.3 |
| | **p-value (B→12)** | | | | | 0.326 | | | | | 0.763 | | | | | 0.833 | | |
| **12th** | **p-value at 12th month** | 47 | ±7.0 | 47.6 | ±7.4 | 0.516 | 46.4 | ±6.6 | 51.7 | ±7.4 | 0.001 | 44.9 | ±5.8 | 45.3 | ±6.3 | 0.247 | 48.3 | ±7.3 |
| **Weight** | | **overall** | **SD** | **OAGB** | **SD** | **p-value** | **RYGB** | **SD** | **male** | **SD** | **p-value** | **female** | **SD** | **T2DM** | **SD** | **p-value** | **nonT2DM** | **SD** |
| **B** | **p-value at baseline** | 130 | ±19 | 127 | ±15 | 0.227 | 135 | ±23 | 138 | ±22 | 0.101 | 127 | ±17 | 132 | ±21 | 0.747 | 130 | ±18 |
| | **p-value (B→6)** | | | | | 0.540 | | | | | 0.922 | | | | | 0.023 | | |
| **6th** | **p-value at 6th month** | 102 | ±16 | 99 | ±14 | 0.120 | 107 | ±19 | 109 | ±18 | 0.185 | 101 | ±16 | 101 | ±16 | 0.386 | 104 | ±17 |
| | **p-value (B→12)** | | | | | 0.236 | | | | | 0.280 | | | | | 0.006 | | |
| **12th** | **p-value at 12th month** | 98 | ±16 | 94 | ±14 | 0.110 | 103 | ±18 | 106 | ±19 | 0.080 | 95 | ±15 | 95 | ±16 | 0.289 | 101 | ±17 |
| **Bmi** | | **overall** | **SD** | **OAGB** | **SD** | **p-value** | **RYGB** | **SD** | **male** | **SD** | **p-value** | **female** | **SD** | **T2DM** | **SD** | **p-value** | **nonT2DM** | **SD** |
| **B** | **p-value at baseline** | 45 | ±6 | 44 | ±6 | 0.399 | 46 | ±6 | 42 | ±5 | 0.052 | 46 | ±6 | 46 | ±6 | 0.166 | 44 | ±6 |
| | **p-value (B→6)** | | | | | 0.950 | | | | | 0.920 | | | | | <0.001 | | |
| **6th** | **p-value at 6th month** | 35 | ±5 | 35 | ±6 | 0.480 | 36 | ±5 | 34 | ±5 | 0.066 | 37 | ±5 | 35 | ±5 | 0.670 | 36 | ±5 |
| | **p-value (B→12)** | | | | | 0.256 | | | | | 0.521 | | | | | <0.001 | | |
| **12th** | **p-value at 12th month** | 34 | ±5 | 33 | ±6 | 0.304 | 35 | ±5 | 33 | ±5 | 0.163 | 35 | ±5 | 34 | ±6 | 0.578 | 35 | ±5 |
| **Hba1c** | | **overall** | **SD** | **OAGB** | **SD** | **p-value** | **RYGB** | **SD** | **male** | **SD** | **p-value** | **female** | **SD** | **T2DM** | **SD** | **p-value** | **nonT2DM** | **SD** |
| **B** | **p-value at baseline** | 39 | ±10 | 38 | ±6 | 0.322 | 41 | ±13 | 38 | ±7 | 0.759 | 40 | ±11 | 47 | ±12 | <0.001 | 34 | ±4 |
| | **p-value (B→6)** | | | | | 0.036 | | | | | 0.792 | | | | | <0.001 | | |
| **6th** | **p-value at 6th month** | 34 | ±5 | 34 | ±3 | 0.484 | 33 | ±6 | 34 | ±4 | 0.197 | 34 | ±5 | 37 | ±4 | 0.002 | 32 | ±4 |
| | **p-value (B→12)** | | | | | 0.112 | | | | | 0.244 | | | | | <0.001 | | |
| **12th** | **p-value at 12th month** | 34 | ±7 | 34 | ±3 | 0.532 | 35 | ±10 | 35 | ±6 | 0.157 | 34 | ±7 | 38 | ±8 | 0.005 | 31 | ±4 |
| **Cholesterol** | | **overall** | **SD** | **OAGB** | **SD** | **p-value** | **RYGB** | **SD** | **male** | **SD** | **p-value** | **female** | **SD** | **T2DM** | **SD** | **p-value** | **nonT2DM** | **SD** |
| **B** | **p-value at baseline** | 4.5 | ±0.9 | 4.5 | ±1.0 | 0.481 | 4.6 | ±0.7 | 4.3 | ±1.3 | 0.178 | 4.6 | ±0.7 | 4.5 | ±1.1 | 0.794 | 4.5 | ±0.8 |
| | **p-value (B→6)** | | | | | 0.592 | | | | | 0.475 | | | | | 0.334 | | |
| **6th** | **p-value at 6th month** | 3.8 | ±0.7 | 3.7 | ±0.7 | 0.163 | 3.9 | ±0.6 | 3.5 | ±0.8 | 0.082 | 3.9 | ±0.6 | 3.7 | ±0.8 | 0.354 | 3.9 | ±0.6 |
| | **p-value (B→12)** | | | | | 0.421 | | | | | 0.924 | | | | | 0.118 | | |
| **12th** | **p-value at 12th month** | 3.9 | ±1.0 | 3.8 | ±0.7 | 0.114 | 4.3 | ±1.2 | 3.8 | ±1.1 | 0.437 | 4.1 | ±0.9 | 3.8 | ±0.9 | 0.361 | 4.1 | ±1.1 |
| **HDL** | | **overall** | **SD** | **OAGB** | **SD** | **p-value** | **RYGB** | **SD** | **male** | **SD** | **p-value** | **female** | **SD** | **T2DM** | **SD** | **p-value** | **nonT2DM** | **SD** |
| **B** | **p-value at baseline** | 1.2 | ±0.2 | 1.2 | ±0.2 | 0.899 | 1.2 | ±0.2 | 1.2 | ±0.2 | 0.426 | 1.3 | ±0.2 | 1.2 | ±0.3 | 0.579 | 1.2 | ±0.2 |
| | **p-value (B→6)** | | | | | 0.681 | | | | | 0.328 | | | | | 0.075 | | |
| **6th** | **p-value at 6th month** | 1.2 | ±0.2 | 1.2 | ±0.2 | 0.221 | 1.3 | ±0.2 | 1.2 | ±0.2 | 0.994 | 1.2 | ±0.2 | 1.1 | ±0.2 | 0.029 | 1.3 | ±0.2 |
| | **p-value (B→12)** | | | | | 0.807 | | | | | 0.037 | | | | | 0.027 | | |
| **12th** | **p-value at 12th month** | 1.4 | ±0.3 | 1.4 | ±0.2 | 0.872 | 1.4 | ±0.3 | 1.4 | ±0.3 | 0.487 | 1.4 | ±0.3 | 1.3 | ±0.3 | 0.021 | 1.5 | ±0.2 |
| **LDL** | | **overall** | **SD** | **OAGB** | **SD** | **p-value** | **RYGB** | **SD** | **male** | **SD** | **p-value** | **female** | **SD** | **T2DM** | **SD** | **p-value** | **nonT2DM** | **SD** |
| **B** | **p-value at baseline** | 2.9 | ±0.8 | 2.8 | ±0.9 | 0.406 | 3 | ±0.7 | 2.8 | ±1.1 | 0.240 | 2.9 | ±0.6 | 2.8 | ±0.9 | 0.345 | 2.9 | ±0.8 |
| | **p-value (B→6)** | | | | | 0.082 | | | | | 0.372 | | | | | 0.637 | | |
| **6th** | **p-value at 6th month** | 2.3 | ±0.7 | 2.2 | ±0.7 | 0.395 | 2.4 | ±0.6 | 2 | ±0.7 | 0.046 | 2.4 | ±0.6 | 2.3 | ±0.7 | 0.786 | 2.3 | ±0.6 |
| | **p-value (B→12)** | | | | | 0.915 | | | | | 0.490 | | | | | 0.851 | | |
| **12th** | **p-value at 12th month** | 2.3 | ±0.8 | 2.2 | ±0.8 | 0.498 | 2.3 | ±0.8 | 2.1 | ±1.1 | 0.387 | 2.3 | ±0.6 | 2.2 | ±0.8 | 0.853 | 2.3 | ±0.7 |
| **Triglycerides** | | **overall** | **SD** | **OAGB** | **SD** | **p-value** | **RYGB** | **SD** | **male** | **SD** | **p-value** | **female** | **SD** | **T2DM** | **SD** | **p-value** | **nonT2DM** | **SD** |
| **B** | **p-value at baseline** | 1.4 | ±0.7 | 1.4 | ±0.6 | 0.678 | 1.4 | ±0.8 | 1.3 | ±0.6 | 0.248 | 1.5 | ±0.7 | 1.7 | ±0.9 | 0.103 | 1.3 | ±0.5 |
| | **p-value (B→6)** | | | | | 0.504 | | | | | 0.608 | | | | | 0.115 | | |
| **6th** | **p-value at 6th month** | 1.1 | ±0.4 | 1.1 | ±0.4 | 0.730 | 1.1 | ±0.3 | 0.9 | ±0.3 | 0.296 | 1.2 | ±0.3 | 1.2 | ±0.4 | 0.112 | 1 | ±0.3 |
| | **p-value (B→12)** | | | | | 0.374 | | | | | 0.311 | | | | | 0.713 | | |
| **12th** | **p-value at 12th month** | 1 | ±0.3 | 0.9 | ±0.3 | 0.371 | 1 | ±0.3 | 0.9 | ±0.3 | 0.246 | 1 | ±0.3 | 1.1 | ±0.3 | 0.050 | 0.9 | ±0.3 |
| **Fasting Glucose** | | **overall** | **SD** | **OAGB** | **SD** | **p-value** | **RYGB** | **SD** | **male** | **SD** | **p-value** | **female** | **SD** | **T2DM** | **SD** | **p-value** | **nonT2DM** | **SD** |
| **B** | **p-value at baseline** | 6.1 | ±1.0 | 5.9 | ±0.9 | 0.526 | 6.3 | ±1.2 | 6.1 | ±0.9 | 0.951 | 6.1 | ±1.1 | 6.8 | ±1.1 | <0.001 | 5.6 | ±0.6 |
| | **p-value (B→6)** | | | | | 0.595 | | | | | 0.211 | | | | | 0.027 | | |
| **6th** | **p-value at 6th month** | 5.4 | ±0.7 | 5.4 | ±0.6 | 0.699 | 5.4 | ±0.8 | 5.7 | ±0.8 | 0.056 | 5.3 | ±0.6 | 5.6 | ±1.0 | 0.091 | 5.3 | ±0.4 |
| | **p-value (B→12)** | | | | | 0.648 | | | | | 0.169 | | | | | 0.053 | | |
| **12th** | **p-value at 12th month** | 5.4 | ±0.9 | 5.4 | ±0.8 | 0.876 | 5.5 | ±1.0 | 5.8 | ±1.2 | 0.042 | 5.3 | ±0.7 | 5.7 | ±1.2 | 0.091 | 5.2 | ±0.5 |
| **Fasting Insulin** | | **overall** | **SD** | **OAGB** | **SD** | **p-value** | **RYGB** | **SD** | **male** | **SD** | **p-value** | **female** | **SD** | **T2DM** | **SD** | **p-value** | **nonT2DM** | **SD** |
| **B** | **p-value at baseline** | 20.3 | ±17.2 | 20.5 | ±14.7 | 0.964 | 20.1 | ±20.0 | 19.1 | ±14.5 | 0.354 | 20.9 | ±18.5 | 24.8 | ±23.6 | 0.943 | 17.2 | ±9.8 |
| | **p-value (B→6)** | | | | | 0.957 | | | | | 0.308 | | | | | 0.631 | | |
| **6th** | **p-value at 6th month** | 8.2 | ±4.4 | 8.8 | ±4.6 | 0.781 | 7.7 | ±4.2 | 8.4 | ±4.6 | 0.863 | 8.2 | ±4.4 | 7.9 | ±3.9 | 0.580 | 8.5 | ±4.8 |
| | **p-value (B→12)** | | | | | 0.700 | | | | | 0.136 | | | | | 0.817 | | |
| **12th** | **p-value at 12th month** | 6.6 | ±4.1 | 7.1 | ±4.6 | 0.647 | 6.2 | ±3.5 | 6.7 | ±3.7 | 0.713 | 6.7 | ±4.3 | 7 | ±4.1 | 0.464 | 6.4 | ±4.1 |
| **Fat kg** | | **overall** | **SD** | **OAGB** | **SD** | **p-value** | **RYGB** | **SD** | **male** | **SD** | **p-value** | **female** | **SD** | **T2DM** | **SD** | **p-value** | **nonT2DM** | **SD** |
| **B** | **p-value at baseline** | 64 | ±13 | 62 | ±12 | 0.549 | 65 | ±15 | 61 | ±16 | 0.244 | 65 | ±12 | 64 | ±12 | 0.790 | 63 | ±14 |
| | **p-value (B→6)** | | | | | 0.645 | | | | | 0.129 | | | | | 0.028 | | |
| **6th** | **p-value at 6th month** | 43 | ±12 | 41 | ±11 | 0.385 | 45 | ±12 | 39 | ±14 | 0.139 | 45 | ±10 | 39 | ±9 | 0.097 | 46 | ±13 |
| | **p-value (B→12)** | | | | | 0.258 | | | | | 0.004 | | | | | 0.016 | | |
| **12th** | **p-value at 12th month** | 38 | ±12 | 36 | ±13 | 0.529 | 39 | ±12 | 31 | ±14 | 0.036 | 40 | ±11 | 35 | ±11 | 0.204 | 40 | ±13 |
| **Fat %** | | **overall** | **SD** | **OAGB** | **SD** | **p-value** | **RYGB** | **SD** | **male** | **SD** | **p-value** | **female** | **SD** | **T2DM** | **SD** | **p-value** | **nonT2DM** | **SD** |
| **B** | **p-value at baseline** | 49 | ±6 | 49 | ±6 | 0.839 | 49 | ±7 | 44 | ±7 | <0.001 | 51 | ±4 | 49 | ±5 | 0.955 | 49 | ±7 |
| | **p-value (B→6)** | | | | | 0.688 | | | | | 0.042 | | | | | 0.182 | | |
| **6th** | **p-value at 6th month** | 41 | ±7 | 41 | ±8 | 0.749 | 41 | ±7 | 35 | ±8 | <0.001 | 43 | ±5 | 39 | ±7 | 0.172 | 42 | ±8 |
| | **p-value (B→12)** | | | | | 0.261 | | | | | <0.001 | | | | | 0.173 | | |
| **12th** | **p-value at 12th month** | 37 | ±9 | 37 | ±10 | 0.790 | 38 | ±9 | 30 | ±9 | <0.001 | 41 | ±6 | 36 | ±8 | 0.333 | 39 | ±9 |
